# Reported fatality ratios and surveillance data readiness: a test applied to health zones in the 2026 Bundibugyo virus disease epidemic, Democratic Republic of the Congo

**DOI:** 10.64898/2026.07.28.26359162

**Authors:** Johan G. L. Verheyden, Celestin Nzanzu Mudogo, Wolfgang Jacquet

**Author notes:** Corresponding author: Johan G. L. Verheyden,. Postal address: Sepal Gardens (Apt. 605), Gatundu Close, Kileleshwa, Nairobi, Kenya.

## Abstract

Dashboards and situation reports publish deaths per confirmed case by health zone, and decisions on investigation and resources lean on these numbers. We ask when such a reported ratio is ready to be read as fatality, and propose a readiness test that combines Bayesian partial pooling, an ascertainment tipping-point analysis and repeated refitting at successive data cut-offs, framed by a surveillance-maturity logic. We applied it to public cumulative confirmed cases and deaths from the 2026 Bundibugyo virus disease epidemic in the Democratic Republic of the Congo (8,728 cases and 4,205 deaths on 6 October). Among 41 health zones with at least 10 cases, North Kivu zones had higher adjusted odds of a reported death than Ituri zones (odds ratio 1.46 (0.99-2.14), 95% credible interval), a point estimate that was insensitive to the prior, the case threshold and omission of single zones. The estimate was nevertheless unstable across cut-offs: the odds ratio was 1.78, 2.54, 1.67, 1.51 and 1.46 on 20 July, 30 August, 30 September, 5 October and 6 October (the last three from identical zones), and its lower credible limit, 1.01 on 5 October, was 0.99 on 6 October. A 1.25-fold difference in the ratio of death to case ascertainment between the provinces would suffice to produce the contrast. Cases and deaths were often reported on the same date, and mapped facility indicators added nothing. By this test, zone-level ratios are not yet interpretable as fatality, although they correctly show where to investigate. In simulated stable epidemics the cumulative decline since 25 August was never matched, whereas the drift ratio over three cut-offs was a weak signal. Surveillance-design actions follow: date and label ratios, show uncertainty and drift, record event dates, audit ascertainment, and link patient records.

## 1. Introduction

### 1.1. The decision problem

During an epidemic, situation reports and dashboards routinely publish the number of deaths per confirmed case, often by health zone. These figures are used to decide where to send investigation teams, how to assign treatment capacity and how to judge whether a response is working. They look simple, but the numerator and the denominator describe different people at different times: recently confirmed patients have not yet had time to die or recover, deaths may be found or confirmed later than the cases they belong to, mild infections may be missed, deaths in the community may be detected unevenly, and reporting systems add cases and deaths in batches (Ghani et al., 2005; Van Kerkhove et al., 2015). The ratio can be observed and is operationally important, but it is not the probability that a newly confirmed person will die. The practical question for surveillance design is therefore not only how to estimate fatality, but when a reported ratio is ready to be read as fatality at all.

### 1.2. Existing approaches and the gap

The statistical literature on case-fatality estimation addresses right-censoring, reporting delays and heterogeneity. Methods correct for outcomes that have not yet occurred (Ghani et al., 2005), for delays between events and their reporting (Bastos et al., 2019; McGough et al., 2020), and for variation by place, time and patient group in West Africa, including imputation and machine-learning approaches (Forna et al., 2020a; Forna et al., 2020b; Forna et al., 2021; Garske et al., 2017). Syntheses of earlier Ebola outbreaks provide benchmarks (Izudi & Bajunirwe, 2024; Rosello et al., 2015; WHO Ebola Response Team, 2014). These methods assume that cases and deaths are ascertained completely, or at a known rate. They do not tell an analyst or a decision-maker whether the surveillance system that produced the counts has reached the state in which such corrections are meaningful. We address that gap: how can an analyst, using only the aggregate data a ministry publishes, judge whether a zone-level ratio is ready to be interpreted?

### 1.3. A readiness framework

We build on a staged framework for data readiness in real-time outbreak response (Verheyden & Nzanzu Mudogo, 2026). It separates two judgments that must precede any reading of a reported ratio as fatality. The first is whether the surveillance system is mature: whether its capacity and processes (testing, case investigation, death recording, reporting) work well enough to give a stable, roughly constant-delay record of real disease activity. Surveillance maturity is a property of capacity and process, shown by independent input and activity evidence, and it cannot be defined by the stability of the data stream it produces. The second is epidemiological maturity: whether the epidemic has generated enough resolved outcomes, over enough time and after clearance of reporting backlogs, for fatality parameters to be identifiable and stable. This presupposes the first. The framework also treats maturity as specific to a reporting channel: the case channel and the death channel need not mature together, and a ratio of the two inherits the difference.

The framework yields a testable expectation. If a zone-level ratio measured a stable property of the epidemic, estimates and comparisons built on it should settle as data accumulate. If it measures an unmatured reporting process, they should keep moving, and they should move in ways that follow changes in what is being reported. We therefore treat the behavior of the ratio across successive cut-offs as evidence and design the analysis to expose it (Table 1).

**Table 1.** How each component of the readiness test relates to the maturity framework.

| Framework question | What would indicate readiness | Component of the test |
| --- | --- | --- |
| Is surveillance mature (independent input and activity evidence)? | Independent evidence of testing, investigation and death-recording performance by zone | Search for such evidence (Section 5.4) |
| Do case and death channels differ in completeness? | Equal ascertainment of cases and deaths ($a_d = a_c$, the fractions of true deaths and cases reported; Section 3.3) | Tipping-point and scenario analysis of $a_d/a_c$ (Section 3.3) |
| Are parameters identifiable and stable (epidemiological maturity)? | Estimates converge as the cut-off moves forward | Refitting at successive cut-offs and a drift ratio (Sections 2.4 and 3.4) |
| Is the report-date record informative about the epidemic process? | Report dates reflect event dates with a stable lag | Same-day reporting and delay diagnostics (Section 2.5) |

*Epidemic age and reporting-date completeness, used here as covariates, are output-side indicators of the data stream and are not measures of maturity in this framework.*

### 1.4. Objectives and contribution

Our objectives were to (i) specify a readiness test for reported fatality ratios that uses only published aggregate data; (ii) apply it to health zones in the 2026 Bundibugyo virus disease (BVD) epidemic in the Democratic Republic of the Congo (DRC); and (iii) translate the result into surveillance-design and policy actions. The contribution is the linkage of data, modeling and policy: a Bayesian model of the reported ratio, an explicit ascertainment tipping point, a stability analysis across data cut-offs, and a mapping from findings to the design of dashboards, audits and data linkage.

### 1.5. The 2026 epidemic

BVD is caused by Bundibugyo virus, first identified in western Uganda in 2007-08 (Wamala et al., 2010). Earlier outbreaks reported lower proportions of deaths than many outbreaks of Ebola virus (MacNeil et al., 2010; Roddy et al., 2012), and the 2012 outbreak in Isiro, DRC, showed substantial mortality and the difficulty of documenting care in the field (Kratz et al., 2015). The 2026 epidemic is much larger, with 2,473 confirmed cases and 999 deaths on 20 July and 8,728 confirmed cases and 4,205 deaths on 6 October 2026 (INRB-UMIE, 2026), mostly in Ituri and North Kivu (Akilimali et al., 2026; Sullivan, 2026; WHO, 2026d; WHO, 2026e). Successive WHO reports document the growth from the first notifications (WHO, 2026a; WHO, 2026b; WHO, 2026c) and the Director-General’s briefing set out the response priorities (WHO, 2026f). The World Health Organization (WHO) has stated that part of the rise reflects expanded surveillance, testing and diagnostic capacity (WHO, 2026d), and has noted that some zones show falling reported cases with rising deaths, which it attributes to differences in ascertainment, reporting delays or lags between detection and death (WHO Regional Office for Africa, 2026). Surveillance in eastern DRC is uneven, insecurity and fragile health services limit case finding and care (ACLED, 2026; Bhadelia, 2026; Doshi et al., 2023; Ilunga Kalenga et al., 2019; Kihanduka et al., 2026; Kyomba et al., 2024; Médecins Sans Frontières, 2026), and the sensitivity of case and death detection is not measured in the situation-report data. This is the central limitation of every analysis of these data, including ours, and we treat it as the main interpretive question.

## 2. Material and methods

### 2.1. Data and definitions

This is an ecological analysis of public aggregate data. The source was the BDBV2026-Data repository of the Institut National de Recherche Biomédicale (INRB) and its Unité de Modélisation et Intelligence Épidémiologique (UMIE), which holds processed national and health-zone series from the situation reports (SitReps) of the Institut National de Santé Publique (INSP) (INRB-UMIE, 2026). The analysis pipeline is pinned to repository commit a3250731ee0ed57ebe977c916aec24670082b33a (8 October 2026, SitRep 145); the data run to 6 October 2026, which is the primary cut-off. The same pipeline was run at 20 July, 10 August, 30 August, 15 September, 30 September and 5 October 2026. No patients were linked to outcomes, so the analysis cannot estimate individual fatality, time to death or competing risks.

The data are manually transcribed from INSP SitReps into the repository. A confirmed case is a person with a positive laboratory test (reverse-transcription polymerase chain reaction, RT-PCR) for Bundibugyo virus, and a confirmed death is a death in such a person; probable and suspected cases and deaths are not included. This is how the authors, together with colleagues involved in the response, interpret the SitRep category; the SitReps and the repository give no formal definition, but SitRep MVE 014 states that suspected deaths were omitted until confirmed, classified as probable or excluded (INSP, 2026). Deaths in the community that were never tested are therefore outside the numerator. National totals come from the national banner of each SitRep; zone totals are the sum of the zone table, and the two do not always agree. Zone names appear under variant spellings and were matched to one standard name using the repository alias table, identically for cases and deaths, before any calculation. Gaps in the series were not filled.

### 2.2. Zone-level descriptors

For zone i, the reported ratio is D_i_/C_i_, where D_i_ is the cumulative number of confirmed deaths and C_i_ the cumulative number of confirmed cases at the cut-off. Jeffreys intervals (a standard interval for a proportion) describe uncertainty due to small numbers only. Three descriptors were used as adjustment variables: epidemic age (days from the zone’s first confirmed case to the cut-off); recent case share (the increase in cumulative cases over the last 14 days divided by the cumulative total); and reporting-date completeness (the proportion of calendar days between the first case and the cut-off on which a numeric cumulative count was reported). These are output-side indicators of the data stream, not measures of surveillance maturity, and reporting-date completeness is not the sensitivity of surveillance.

### 2.3. Design of the readiness test

The test has four components, each addressing a question in Table 1: (a) a model of the reported ratio with partial pooling across zones and a province contrast (Section 3.1); (b) an ascertainment tipping-point analysis that asks how large a difference between case and death reporting would be needed to produce the contrast without any real difference in outcome (Section 3.3); (c) the same model refitted at successive data cut-offs, summarized by a drift ratio (Sections 2.4 and 3.4); and (d) diagnostics of how cases and deaths are reported in time (Section 2.5). The model was fitted to the zones with at least 10 confirmed cases, and descriptive tables foreground zones with at least 20.

### 2.4. Robustness and stability analyses

We (i) repeated the fit with coefficient prior standard deviations of 1, 2.5 and 5; (ii) refitted with minimum case thresholds of 1, 3, 5, 10 and 20; (iii) refitted 41 times omitting one zone each time and recorded the range of the North Kivu odds ratio and the number of fits whose interval excluded 1; (iv) examined predictor correlations and variance inflation factors (values near 1 indicate that predictors are not redundant); and (v) repeated the primary model at seven cut-offs (20 July, 10 August, 30 August, 15 September, 30 September, 5 October and 6 October) with an unchanged specification. Item (v) is the most direct test of whether the estimates describe a stable feature of the epidemic or a snapshot.

### 2.5. Secondary and exploratory analyses

Same-day reporting. Daily increments in cumulative cases and deaths were calculated only between consecutive calendar days with numeric values; negative increments (administrative revisions) were excluded from correlations but counted. Spearman correlations between case and death increments were computed at lags 0 to 14 days (at least 8 paired observations). We report the number of zones whose strongest association was at lag 0, the median lag-0 correlation, and the median share of dates with new deaths that also had new cases, among zones with at least 20 cases and at least three positive death-increment days. Delay diagnostic. A negative-binomial model of daily national deaths, driven by earlier case increments through a kernel with a same-day mass and a gamma distribution over lags 1 to 21 days, was fitted with and without a reporting-shock term; details are in Appendix C.

Facility indicators. Static pre-outbreak inventories were used as exploratory proxies: the Humanitarian OpenStreetMap Team (HOT) health-facility layer for the DRC, distributed through the Humanitarian Data Exchange (OpenStreetMap snapshot of 5 October 2026; 5,252 features; ODbL license) and GRID3 (Geo-Referenced Infrastructure and Demographic Data for Development) health-site counts (reference year 2025) and Healthsites.io counts carried in the health-zone polygon file of the repository (extraction date not recorded). These layers show where facilities were mapped before the epidemic, not which were open, staffed, supplied or used. An earlier snapshot (19 June 2026; 5,303 features) was used for the first analysis and gave the same conclusions (Appendix A). Two indicators (clinical facilities per 100,000 population; distance from the zone’s representative point to the nearest mapped hospital) were added one at a time to the primary model and compared by the small-sample corrected Akaike information criterion (AICc; lower is better) and by leave-one-out predictive log score (higher is better), with the paired difference from the primary model and its standard error. Appendix B lists every candidate predictor examined and its status relative to the data.

### 2.6. Software, reproducibility and ethics

The analyses are scripted end to end (Python 3.13), one script per step, with fixed seeds. Each step run at the 30 August 2026 cut-off reproduces the zone, same-day reporting and delay outputs of the earlier analysis exactly, and the model fits to within Monte Carlo error. A random sample of the transcribed zone values was checked against the source PDFs (Section 2.7). The primary model was fitted by an adaptive random-walk Metropolis sampler (four chains, 2,000 warm-up and 25,000 retained draws per chain) and checked at the 6 October cut-off by an independent fit with the No-U-Turn sampler (NumPyro), with agreement to within 0.02 on the odds-ratio scale and across further seeds and cut-offs (Appendix F). Other fits used shorter runs: leave-one-zone-out fits two chains of 4,000 draws, the weekly refits of the real series 15,000 draws per chain and the simulated epidemics 4,000 draws per chain, all with 1,000 to 2,000 warm-up draws, so values for the same cut-off can differ in the second decimal between tables. The 5 October outputs were regenerated from the later repository commit and were identical to those obtained from the commit available on 5 October.

Ethics. The study used only public, aggregate, non-identifiable secondary data, so no ethics approval or individual consent was required. The authors’ wider program of research on the 2026 BVD epidemic is covered by approval UNILU/CEM/089/2026 of the Medical Ethics and Research Committee of theUniversity of Lubumbashi (Comité d’Éthique Médicale et de la Recherche, CEM-UNILU), dated 5 October 2026.

### 2.7. Transcription check

Because the repository holds hand-transcribed values, one author-independent check was made: for five SitReps (093, 108, 125, 140 and 144; 15 August to 5 October 2026), every health-zone cumulative count of confirmed cases and deaths and the national totals were compared with the PDF tables, 633 comparisons in all, and all matched exactly. SitReps earlier than 093 could not be checked because their PDFs were not retrievable from the repository’s large-file storage, and the latest report (SitRep 145, the 6 October data) could not be checked for the same reason. In the PDFs, deaths not yet assigned to a zone appear as a separate row, which explains the difference between national and zone death totals.

## 3. Theory/calculation

### 3.1. Beta-binomial model of the reported ratio

Estimand. The model describes the proportion of confirmed cases that appear as reported deaths in each zone at the cut-off, and how it differs between provinces after adjustment. It does not estimate the probability that a patient dies. For zone i with n_i_ confirmed cases and d_i_ reported deaths,

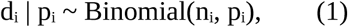

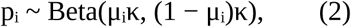

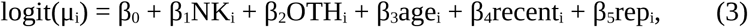

where NK_i_ and OTH_i_ indicate North Kivu and the other provinces (Haut-Uélé and Tshopo in the modeled set; Ituri is the reference), and age_i_, recent_i_ and rep_i_ are the standardized epidemic age, recent case share and reporting-date completeness. The concentration κ governs how much zones vary around their regression mean beyond binomial variation. Priors were Normal(0, 2.5) for each β_j_ and log κ ∼ Normal(log 20, 1.5). Coefficients are reported as posterior medians of exp(β_j_), that is, odds ratios (OR): an OR of 1.5 means that the odds of a reported death among confirmed cases are 1.5 times higher than in the reference, other things equal. The 95% credible interval (CrI) is the Bayesian counterpart of a confidence interval. Appendix G is a glossary of the statistical terms.

### 3.2. Partially pooled zone estimates

The partially pooled value for zone i is the posterior of its latent proportion,

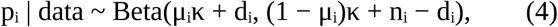

which lies between the zone’s own ratio and its regression mean; the weight on the regression mean is κ/(κ + n_i_). These estimates stabilize reported ratios. They do not correct for unresolved outcomes or for missed cases or deaths.

### 3.3. Ascertainment and the tipping point

Suppose a fraction a_c_ of true cases and a fraction a_d_ of true deaths are reported. The reported ratio is then approximately

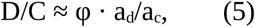

where φ is the true proportion. A reported ratio D/C is feasible only if a_d_/a_c_ ≥ D/C, and a difference in the reported ratio between provinces can be produced entirely by a difference in a_d_/a_c_. At average covariate values we computed

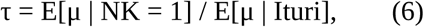

the ratio of the modeled mean reported proportion in North Kivu to that in Ituri. Under Equation 5, τ is the multiplicative difference in a_d_/a_c_ that would, with no real difference in outcome, produce the estimated contrast (the tipping point). We report its posterior median and 95% credible interval and the probabilities that τ exceeds 1.25 and 1.5. We also tabulated the implied fatality (D/C)(a_c_/a_d_) over case ascertainment 0.2 to 1.0 and death ascertainment 0.1 to 1.0. These are scenario calculations, not estimates of sensitivity: they show how large a difference in ascertainment would matter, not whether it exists.

### 3.4. Drift ratio across cut-offs

To summarize stability with one number we define, for a cut-off t and the three most recent cut-offs W(t) up to and including t,

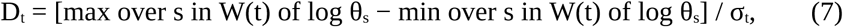

where θ_s_ is the North Kivu odds ratio fitted at cut-off s and σ_t_ = [log(upper limit) − log(lower limit)]/(2 × 1.96) is the posterior standard deviation on the log scale at t. A drift ratio below 1 means that the estimate moved by less than one posterior standard deviation over the window; a ratio above 1 means that it moved by more than its own sampling uncertainty. The drift ratio is a descriptive device, and we calibrate it by simulation (below).

Calibration. To learn what drift to expect when nothing changes, we simulated stable epidemics. True zone-level proportions were drawn once per simulated epidemic from the model fitted at 6 October (true North Kivu odds ratio 1.43 and 1.36 on average in the two scenarios, with the observed covariates and case series), and each case died with its zone’s proportion, either on the day of confirmation (no delay) or after a gamma-distributed delay with mean 7 days. The primary model was refitted at 12 weekly cut-offs (21 July to 6 October) in each of 20 simulated epidemics per scenario, and the drift ratio was computed over windows of three successive weekly cut-offs. In the real data the same weekly refits were made in three phases (cut-offs ending 6 October, 3 October and 30 September), giving 28 windows, and we related each window’s drift ratio to the absolute change over the following week, in posterior standard deviations (Spearman correlation). We also computed the cumulative standardized change from a reference cut-off r to the final cut-off f,

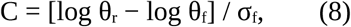

with r = 25 August and f = 6 October, and compared it with the same statistic in the simulated stable epidemics.

## 4. Results

### 4.1. Data coverage and reconciliation

At 6 October, 65 zones had a numeric case count; 64 had at least one confirmed case, 41 at least ten and 35 at least twenty. The zone tables summed to 8,728 cases and 3,712 deaths (42.5%); the national banner gave 8,728 cases and 4,205 deaths (48.2%). Cases agreed exactly, and the zone table contained 493 fewer deaths than the national banner (Table A.1). Differences between national and zone totals are routine in the SitReps. WHO reported on 23 August that 250 cumulative deaths had not yet been assigned to a health zone (WHO Regional Office for Africa, 2026), which is consistent with this explanation, although we cannot establish that they are the same deaths. The national banner was used for national figures and the zone table for geographic comparisons; zone-level conclusions assume that the unassigned deaths are not concentrated in the zones compared. We found no zone-level evidence of testing capacity, case investigation or death-recording performance for the period, because the repository’s zone series for suspected cases and contacts end on 30 May 2026 and its conflict data are provincial and end in the same month; this absence is itself a finding for the readiness assessment (Section 5.4).

### 4.2. The national ratio rose, then flattened

The national ratio rose from 17.4% on 1 June to 31.0% on 1 July, 40.4% on 20 July and 47.0% on 15 August, and has been flat since: 48.4% on 30 August, 48.3% on 15 September, 48.4% on 30 September and 48.2% on 6 October (Fig. 1). This plateau is what would be expected if the right-censoring of recent cases has largely worked through at national level; it does not show that reporting is complete. Early values were volatile: on 28 May confirmed cases rose from 125 to 210 while deaths stayed at 17, lowering the ratio from 13.6% to 8.1%, which the SitRep attributed to laboratory processing and reclassification of suspected cases (INSP, 2026).

**Fig. 1.**
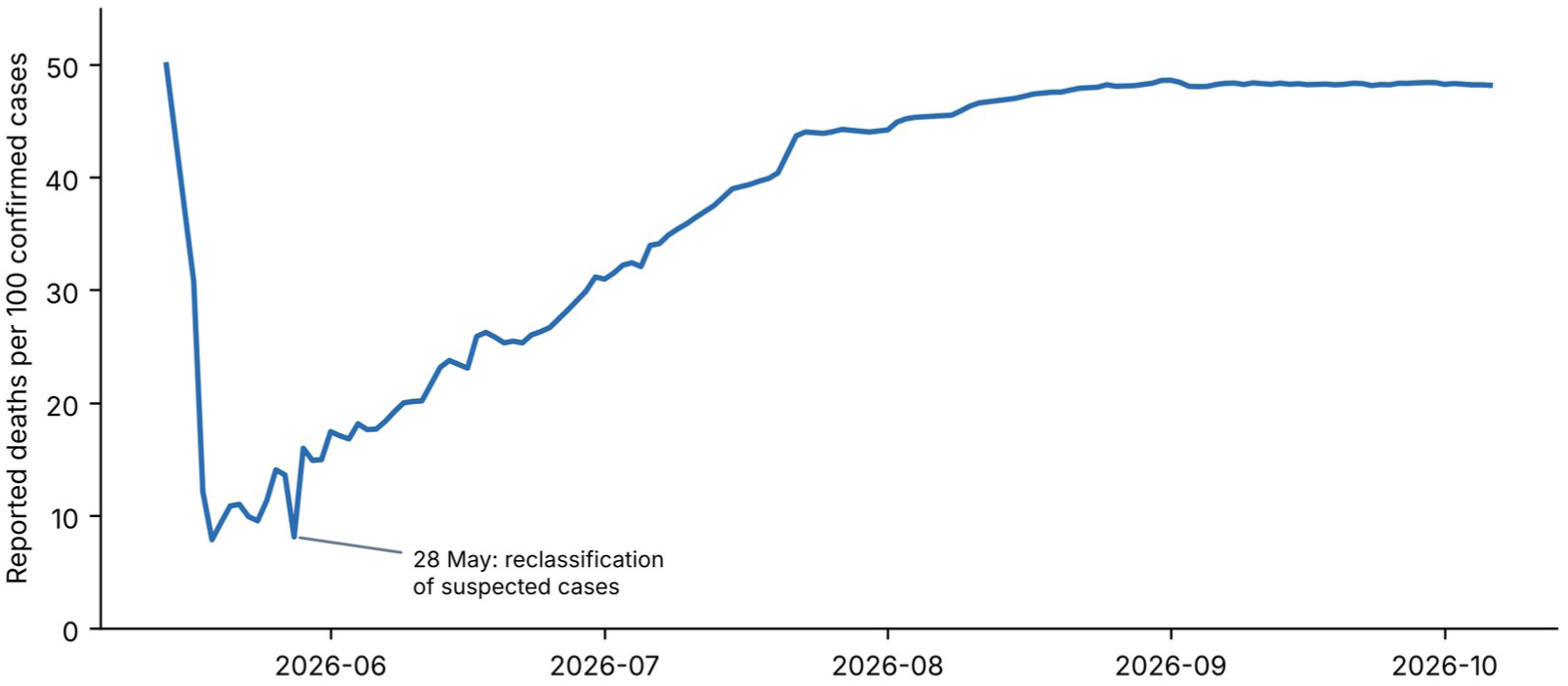
National reported ratio of deaths to confirmed cases over time.

### 4.3. Variation between health zones

Among 35 zones with at least 20 cases, the reported ratio ranged from 20.0% (Mutwanga) to 65.5% (Beni) (Fig. 2; Table A.2). The spread was not due to small numbers: the 11 zones with more than 200 cases ranged from 22.4% (Bambu) to 65.5% (Beni). The three highest ratios were in Beni, Butembo and Katwa (North Kivu), followed by Komanda and Nia Nia (Ituri); the median ratio among these zones was 47% in North Kivu and 36% in Ituri. The concentration parameter κ had a posterior median of about 22 (95% CrI 12 to 39), so a zone with 20 cases takes about half its pooled estimate from the regression and a zone with 500 cases about 4%. Partial pooling moved small zones toward their regression mean but left the ordering largely unchanged (Fig. A.1). North Kivu was not uniformly high: several newer North Kivu zones were low (Mabalako 39.2%, Masereka 32.4%, Kalunguta 34.9%, Vuhovi 28.6%).

**Fig. 2.**
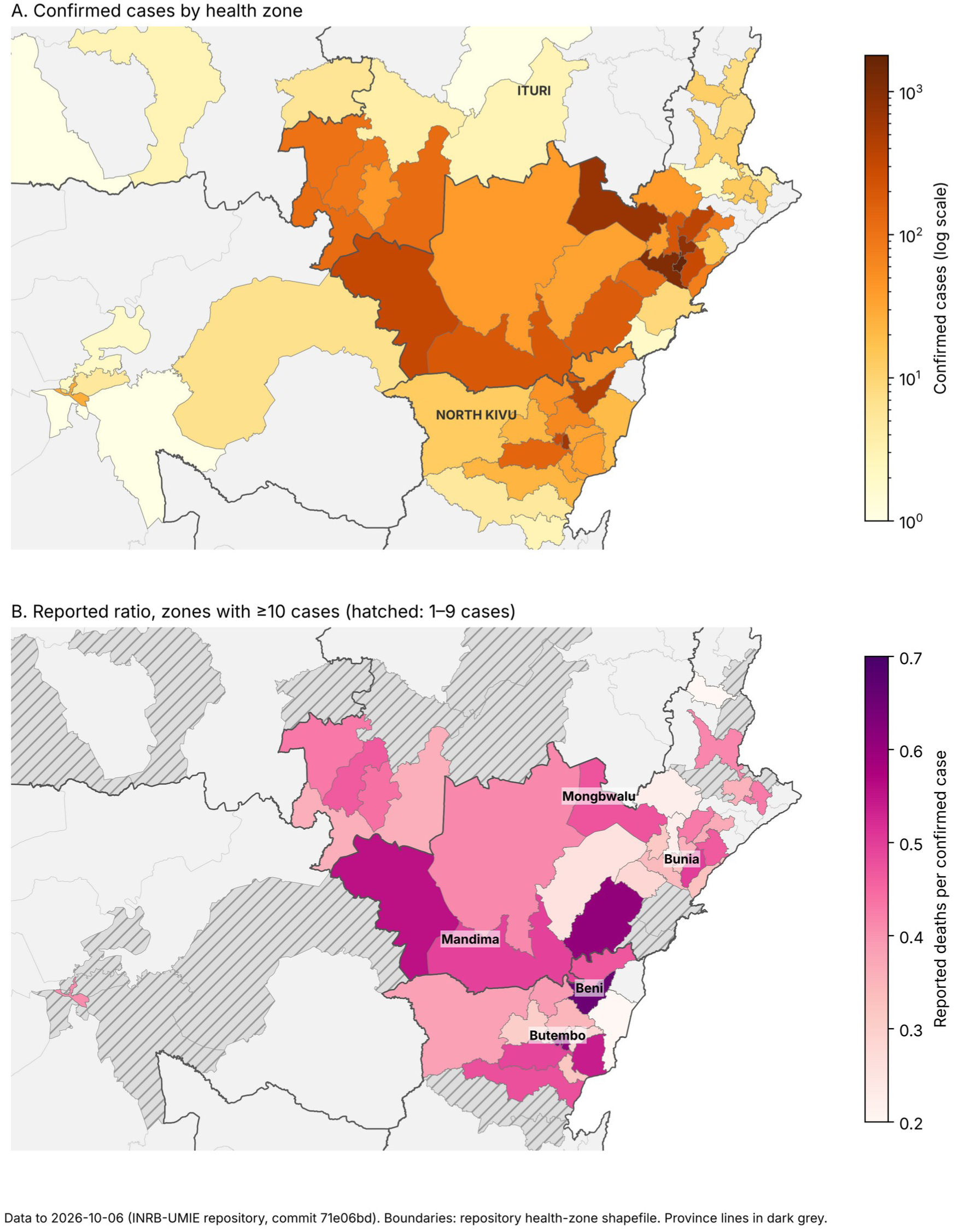
Confirmed cases (A) and reported deaths-to-confirmed-cases ratio (B) by health zone, 6 October 2026. Dark lines are province boundaries. Zones with fewer than 10 cases are hatched in panel B.

### 4.4. The North Kivu-Ituri difference

Ituri accounted for most of the burden (6,513 cases, 2,514 deaths, 38.6%); North Kivu had 1,785 cases and 1,021 deaths (57.2%, Jeffreys interval 54.9%-59.5%); Haut-Uélé had 364 cases (42.0%). In the primary model North Kivu had higher adjusted odds of a reported death than Ituri (OR 1.46 (0.99-2.14); probability that OR > 1, 0.973; Table 2). Epidemic age was positively associated (1.29 (1.04-1.62)); recent case share (1.08 (0.89-1.29)) and reporting-date completeness (0.91 (0.72-1.14)) were not. Epidemic age and reporting-date completeness are no longer collinear (r = 0.28; all variance inflation factors below 2.4), in contrast with 30 August (r = 0.93).

**Table 2.** Primary beta-binomial model, 6 October 2026 (41 zones).

| Term | Posterior OR | 95% CrI | Pr(OR > 1) |
| --- | --- | --- | --- |
| North Kivu vs Ituri | 1.46 | 0.99-2.14 | 0.973 |
| Other provinces vs Ituri | 1.58 | 0.90-2.83 | 0.943 |
| Epidemic age (per SD) | 1.29 | 1.04-1.62 | 0.989 |
| Recent 14-day case share (per SD) | 1.08 | 0.89-1.29 | 0.786 |
| Reporting-date completeness (per SD) | 0.91 | 0.72-1.14 | 0.189 |

*OR, odds ratio; CrI, credible interval; SD, standard deviation.*

### 4.5. Robustness at the 6 October cut-off

Changing the prior standard deviation from 2.5 to 1 or 5 gave 1.43 (0.97-2.07) and 1.46 (1.00-2.16). Refitting with minimum case thresholds of 1, 3, 5, 10 and 20 gave 1.42, 1.43, 1.45, 1.45 and 1.46 (64 to 35 zones); the lower interval limit ranged from 0.96 to 0.98, so that at every threshold the lower limit was at or within a few hundredths of 1. Omitting each zone in turn gave 1.33 to 1.67, with the interval excluding 1 in 15 of 41 fits, compared with 24 of 41 on 5 October and 40 of 41 on 30 September (Fig. A.2, Table D.1); the most influential zone was Mandima. The coefficient for epidemic age was less stable: its interval excluded 1 in 39 of 41 leave-one-out fits and was 1.21 (0.95-1.56) when only zones with at least 20 cases were used. The point estimate was therefore robust to analytic choices at this cut-off, but whether its interval cleared 1 was not.

### 4.6. Stability across cut-offs

The same model fitted at seven cut-offs gave North Kivu odds ratios of 1.78, 2.38, 2.54, 2.09, 1.67, 1.51 and 1.46, and epidemic-age odds ratios of 1.43, 0.72, 0.98, 1.16, 1.26, 1.26 and 1.29 (Table 3, Figs. 3-5). The interval for North Kivu included 1 on 20 July and excluded it at every later cut-off until 6 October, when its lower limit was 0.99. The point estimate rose and then fell as the number of North Kivu zones entering the model increased from 5 to 14. The three most recent cut-offs are the most telling: they use the same 41 zones, yet within six days the odds ratio fell from 1.67 (1.12-2.51) to 1.51 (1.01-2.20) and 1.46 (0.99-2.14), and the number of leave-one-zone-out fits whose interval excluded 1 fell from 40 to 24 to 15 of 41. The tipping point moved in step, from 1.34 to 1.25. The drift ratio (Equation 7) exceeded 1 in 3 of the 5 windows that could be computed (1.39, 0.63, 2.02, 1.67, 0.70); because the cut-offs in these windows are unevenly spaced, Section 4.7 recomputes it at regular intervals. The sign of the epidemic-age coefficient changed twice. The crude provincial ratio in North Kivu behaved similarly (67.7% on 30 August, 59.3% on 30 September, 57.7% on 5 October and 57.2% on 6 October). Conclusions drawn at any single cut-off about either coefficient would therefore have been different at another, and a cut-off a week later will probably differ again.

**Fig. 3.**
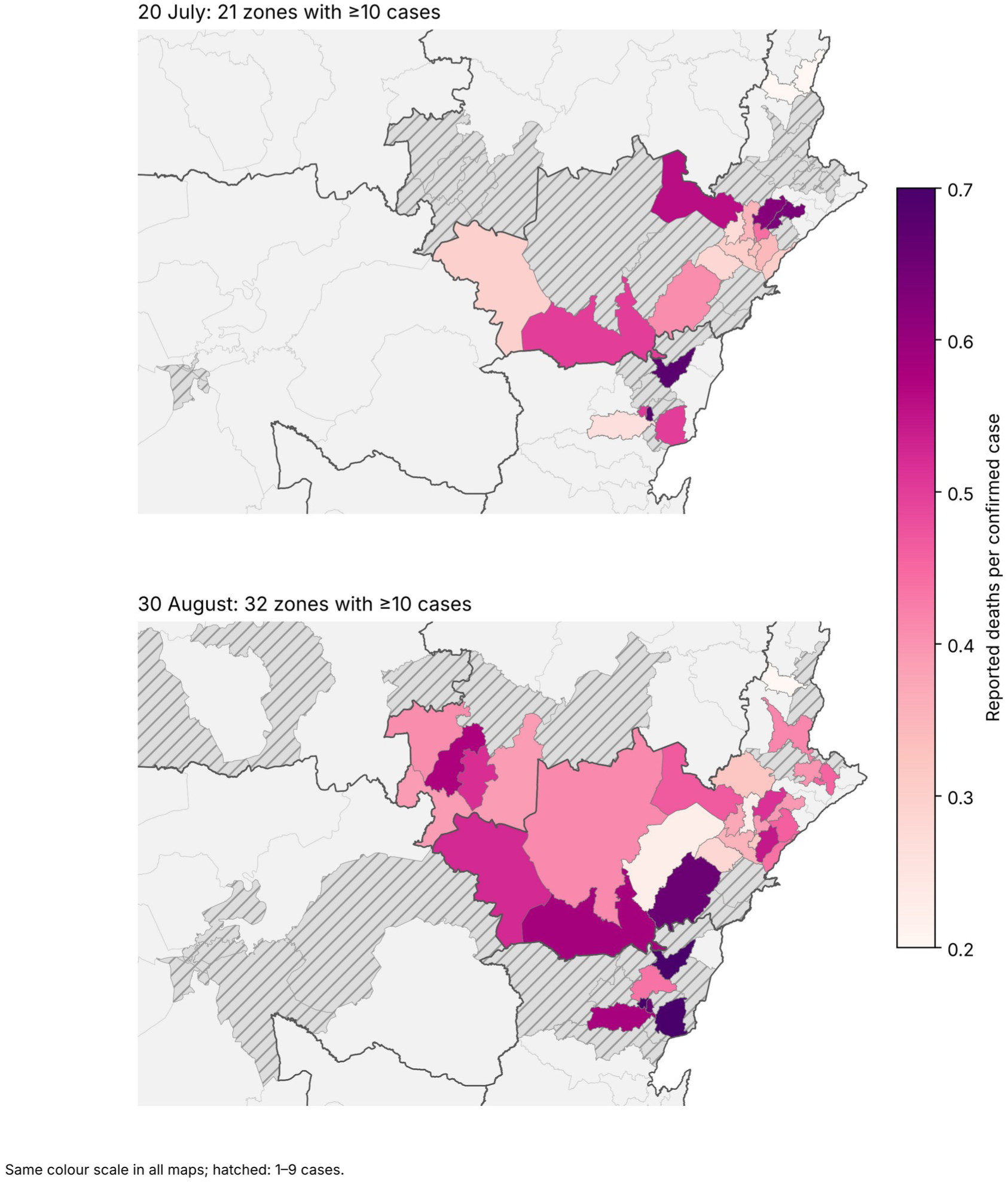
Reported ratio by health zone on 20 July (top) and 30 August 2026 (bottom), on one color scale. Hatched zones have 1 to 9 cases.

**Fig. 4.**
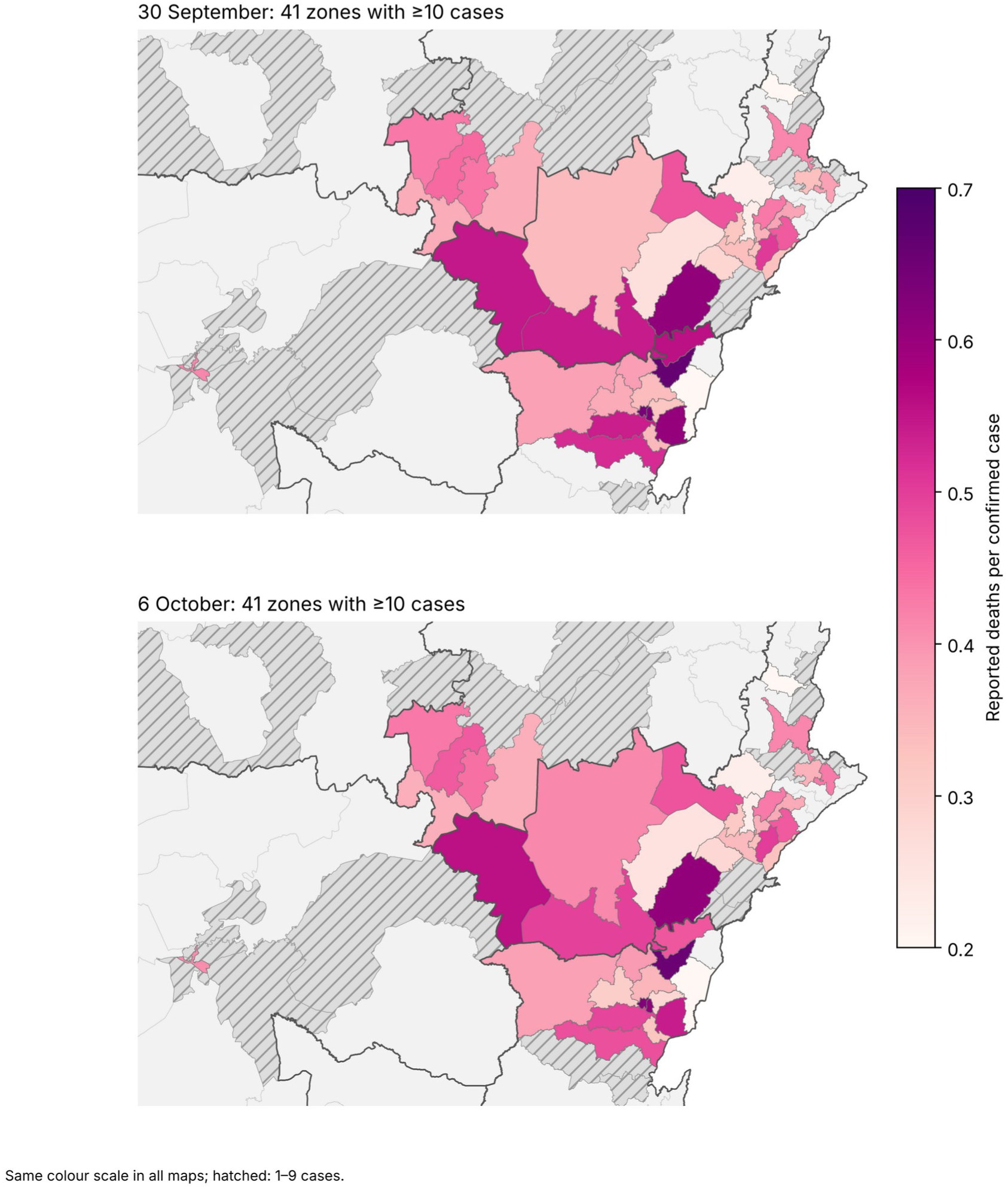
As Fig. 3, on 30 September (top) and 6 October 2026 (bottom). The number of mappable zones and many ratios change from one cut-off to the next.

**Fig. 5.**
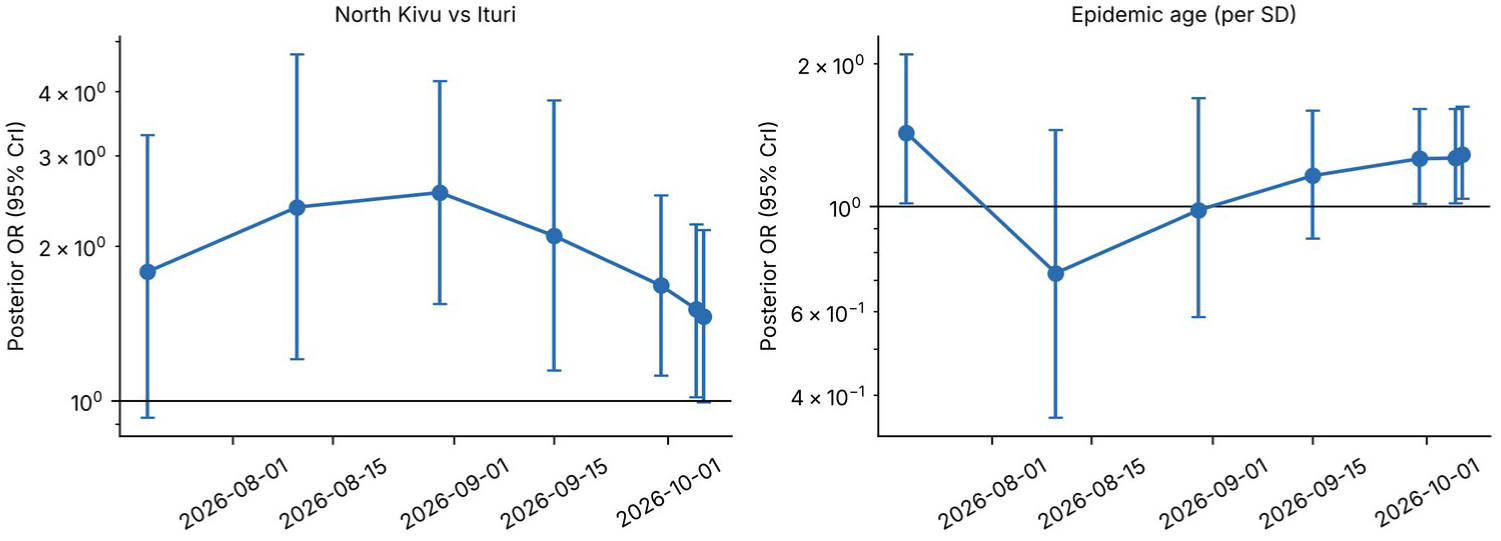
North Kivu odds ratio and epidemic-age odds ratio (95% credible intervals) by cut-off.

**Table 3.**
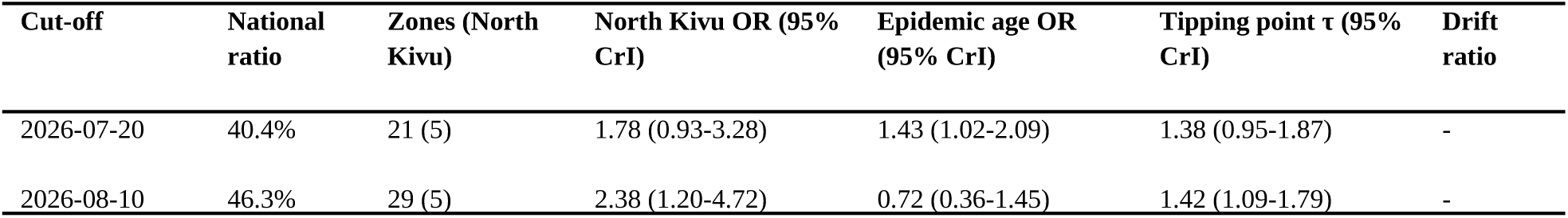
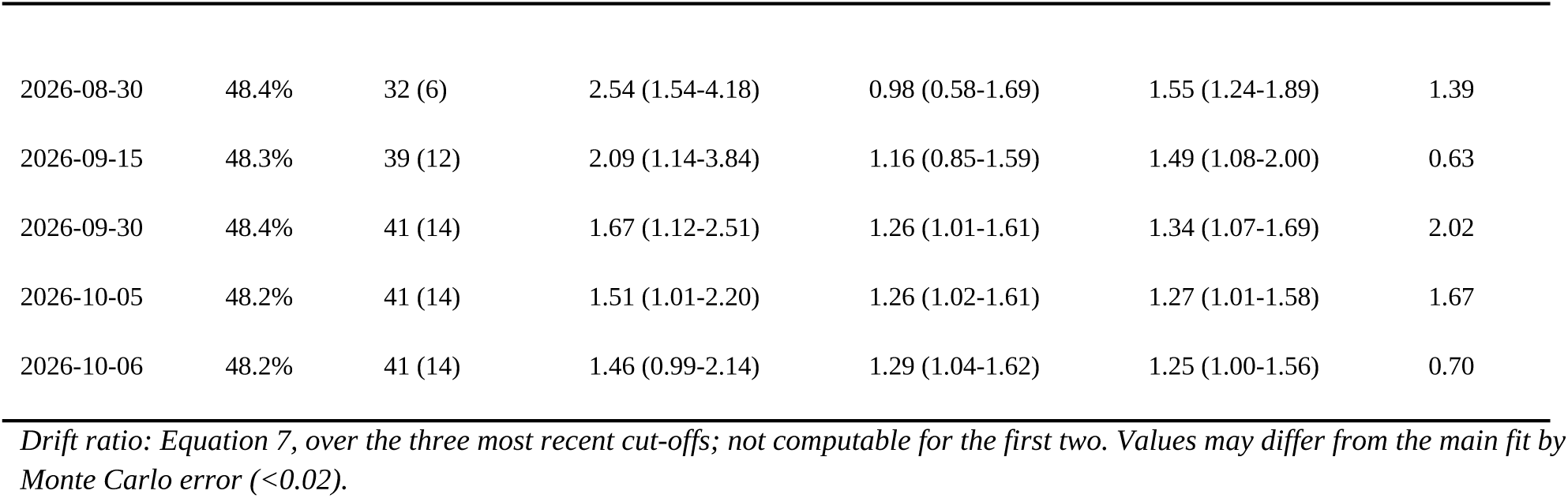
Stability of the primary model across cut-offs (same specification; the 30 September, 5 October and 6 October rows use the same 41 zones).

| Cut-off | National ratio | Zones (North Kivu) | North Kivu OR (95% CrI) | Epidemic age OR (95% CrI) | Tipping point $\tau$ (95% CrI) | Drift ratio |
| --- | --- | --- | --- | --- | --- | --- |
| 2026-07-20 | 40.4% | 21 (5) | 1.78 (0.93-3.28) | 1.43 (1.02-2.09) | 1.38 (0.95-1.87) | - |
| 2026-08-10 | 46.3% | 29 (5) | 2.38 (1.20-4.72) | 0.72 (0.36-1.45) | 1.42 (1.09-1.79) | - |
| 2026-08-30 | 48.4% | 32 (6) | 2.54 (1.54-4.18) | 0.98 (0.58-1.69) | 1.55 (1.24-1.89) | 1.39 |
| 2026-09-15 | 48.3% | 39 (12) | 2.09 (1.14-3.84) | 1.16 (0.85-1.59) | 1.49 (1.08-2.00) | 0.63 |
| 2026-09-30 | 48.4% | 41 (14) | 1.67 (1.12-2.51) | 1.26 (1.01-1.61) | 1.34 (1.07-1.69) | 2.02 |
| 2026-10-05 | 48.2% | 41 (14) | 1.51 (1.01-2.20) | 1.26 (1.02-1.61) | 1.27 (1.01-1.58) | 1.67 |
| 2026-10-06 | 48.2% | 41 (14) | 1.46 (0.99-2.14) | 1.29 (1.04-1.62) | 1.25 (1.00-1.56) | 0.70 |

*Drift ratio: Equation 7, over the three most recent cut-offs; not computable for the first two. Values may differ from the main fit by Monte Carlo error (<0.02).*

### 4.7. Calibration of the drift ratio

In 20 simulated stable epidemics with deaths accruing on the day of confirmation, the drift ratio had a median of 0.47 and a 95th percentile of 1.37, and exceeded 1 in 12% of windows; with a mean delay of 7 days the median was 0.69, the 95th percentile 1.60 and 27% of windows exceeded 1 (Table 4). In the real weekly series the median was 0.81, 9 of 28 windows (32%) exceeded 1 and 2 exceeded 1.6 (those ending on 8 September and 6 October). The windowed drift ratio did not predict the movement of the following week (Spearman rho = -0.15, n = 25, P = 0.47). Over three successive cut-offs the drift ratio is therefore a weak signal: a threshold of 1 would flag roughly one window in eight in a stable epidemic without delay and one in four with delay. The cumulative standardized change discriminated much better. The North Kivu odds ratio fell from 2.81 on 25 August to 1.46 on 6 October, 3.2 posterior standard deviations of the final fit, whereas none of the 20 simulated stable epidemics in either scenario produced a change that large (maxima 2.1 and 1.9; 95th percentiles 2.0 and 1.5). The reference cut-off was chosen after looking at the series, close to its peak, so this comparison is illustrative; taking 4 August instead, the change was 2.6 standard deviations, matched or exceeded in 1 of 20 simulated epidemics without delay and 0 of 20 with delay. With 20 replicates per scenario this is a reference distribution, not a significance test.

**Table 4.** Drift ratio and cumulative standardized change: real series and simulated stable epidemics.

| Statistic | Real series | Stable, no delay | Stable, 7-day delay |
| --- | --- | --- | --- |
| Windows (weekly cut-offs) | 28 | 200 | 200 |
| Drift ratio, median | 0.81 | 0.47 | 0.69 |
| Drift ratio, 95th percentile | - | 1.37 | 1.60 |
| Share of windows with drift ratio above 1 | 32% | 12% | 27% |
| Cumulative standardized change, 25 August to 6 October | 3.2 | median -0.1; 95th percentile 2.0; maximum 2.1 | median 0.3; 95th percentile 1.5; maximum 1.9 |
| Simulated epidemics at least as extreme as the real change | - | 0 of 20 | 0 of 20 |

*Real series: weekly refits in three phases (Appendix H). Simulations: 12 weekly cut-offs, true North Kivu odds ratio about 1.4, no change in surveillance.*

### 4.8. Could differences in surveillance alone produce the contrast?

At average covariate values, the modeled reported proportion in North Kivu is 1.25 times (95% CrI 1.00-1.56) that in Ituri (τ, Equation 6); the ratio of crude provincial ratios is 1.48. The adjusted contrast is therefore equivalent to a 1.25-fold difference in the ratio of death ascertainment to case ascertainment between the provinces (1.39 and 1.39 in models with facility indicators). The probability that τ exceeds 1.25 is 0.49 and that it exceeds 1.5 is 0.05 (Fig. 6). If deaths, relative to cases, were found and reported about a quarter more completely in North Kivu than in Ituri, no real difference in outcome would be needed to explain the data. This does not show that such a difference exists. The observed national ratio requires a_d_/a_c_ of at least 0.48, which rules out some combinations of case and death ascertainment but not most (Fig. A.3). Under the original grid (case ascertainment 0.3 to 0.9, death ascertainment 0.8 to 1.0) the implied fatality ranged from 14.5% to 54.2%; widening the grid to 0.2-1.0 and 0.1-1.0 gives 9.6% to 96.4% among feasible cells. The width of the range is created by the assumptions, not by the data.

**Fig. 6.**
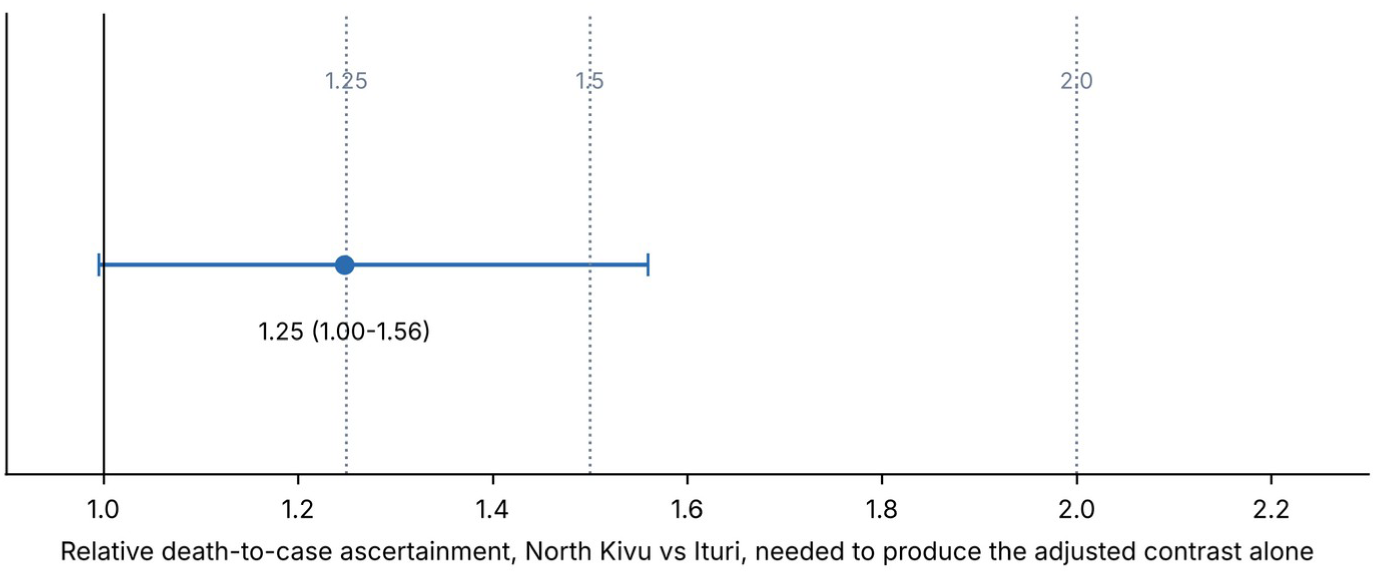
Relative death-to-case ascertainment, North Kivu versus Ituri, needed to produce the adjusted contrast with no real difference in outcome.

### 4.9. Reporting patterns and facility indicators

Among 48 zones with estimable lag correlations, 34 had their strongest association at lag 0. In the 33 zones with at least 20 cases and three or more positive death-increment days, 25 peaked at lag 0; the median lag-0 Spearman correlation was 0.57, and on a median 93.3% of dates with new deaths new cases were also reported. There were 19 zone-days with downward revisions of cases and 8 of deaths. Report dates therefore often reflect when records were submitted, confirmed or reconciled, not the course of illness. The delay model could not separate same-day reporting from positive lags: the free fit placed 63% of mass on the same day with the positive-lag mean at the upper search bound (20 days), and fixed kernels with very different meanings fitted comparably (Appendix C). No clinical delay can be read from these data. Mapped facility indicators were not associated with the reported ratio: after adjustment, clinical facilities per 100,000 had an OR of 0.86 (0.68-1.08) per SD and distance to the nearest mapped hospital 1.14 (0.91-1.42) (Table A.3), and neither improved prediction over the primary model.

## 5. Discussion

### 5.1. Principal findings

Applied to health zones in the 2026 BVD epidemic, the readiness test gave a clear answer. The reported ratio varied widely between zones (20% to 66% among zones with at least 20 cases), and a North Kivu-Ituri difference persisted in the point estimate after adjustment, across priors, case thresholds and omission of any single zone, although by 6 October its credible interval only just reached 1. The difference also moved from one cut-off to the next, a difference of about a quarter in how completely deaths are found relative to cases would be enough to create it, cases and deaths were commonly reported together, and no independent zone-level evidence of surveillance performance was available. The ratio is therefore an administrative quantity that identifies where to look, not a measure of fatality or of the quality of care.

### 5.2. Instability as the finding

The estimates moved with each cut-off, and they will move again. Each cut-off added zones, shifted the share of recent cases and changed which North Kivu zones carried weight; the odds ratio and the epidemic-age coefficient followed. Between 30 September and 6 October the zones were identical and the odds ratio still fell from 1.67 to 1.46, with the lower credible limit passing from 1.12 to 0.99, so accumulating cases and deaths, late confirmations and reclassifications alone were enough to change the answer. A property of patients, such as the risk of dying, would not behave like this once enough data are in; a property of an evolving reporting process would. We therefore read the instability as evidence on the first question of the framework: the zone-level ratio has not reached the stability expected of a quantity that can be interpreted as fatality. Estimates published at any single date, including ours, should be read as dated snapshots, and the date should be part of the claim. This is also why we did not seek the "right" cut-off: the primary analysis is at 6 October because it is the latest available, and the other cut-offs are results, not sensitivity checks.

### 5.3. What the North Kivu-Ituri contrast may and may not mean

The pattern is compatible with at least three explanations that aggregate data cannot separate: differences in the patients and their care, which in Ebola disease depend on factors such as viral load at admission, supportive care and therapeutics (Bah et al., 2015; Fitzpatrick et al., 2015; Lamontagne et al., 2018; Mulangu et al., 2019; Skrable et al., 2017); differences in who is confirmed, for example if more severe cases are confirmed in North Kivu; and differences in how completely deaths are found relative to cases. The tipping-point analysis shows that the third does not need to be large: about a 1.25-fold difference suffices. A qualitative appraisal of the conflict context is in Appendix E. Insecurity, disrupted health services and community distrust, which have shaped earlier epidemics in the region (ACLED, 2026; Doshi et al., 2023; Ilunga Kalenga et al., 2019; Kyomba et al., 2024; Médecins Sans Frontières, 2026), could act through any of the three. WHO reports that surveillance performance differs by province, for example alert investigation reached 67% in Ituri compared with 98-100% in North Kivu, Tshopo and

Haut-Uélé, and that 56.6% of deaths in the week to 23 August occurred in the community (WHO Regional Office for Africa, 2026). Alert investigation concerns case finding, and weaker case finding in Ituri would raise Ituri’s ratio, the opposite of the observed contrast, so it does not explain the finding; it shows only that province-level differences in surveillance performance exist and are documented. Other analyses of the same situation reports have addressed geographic expansion and growth projections (Lupande Mwenebitu et al., 2026), not zone-level reporting ratios. A field assessment of case and death ascertainment in the zones concerned would settle much that these data cannot.

Early in an epidemic, ratios are low because many recent cases have not yet had an outcome, and this should wane as the epidemic matures. The national ratio has been flat near 48% for more than a month, consistent with that. At zone level, however, the epidemic-age coefficient was positive on 20 July, null on 10 August, 30 August and 15 September, positive again on 30 September, 5 October and 6 October, and null when only zones with at least 20 cases were used. We make no claim about it beyond noting that it is a fragile, cut-off-dependent coefficient in a model whose adjustment variables are output-side indicators.

### 5.4. Readiness: what the data can and cannot support

In the framework of surveillance and epidemiological maturity (Verheyden & Nzanzu Mudogo, 2026), reading a zone ratio as a fatality quantity requires both conditions. For surveillance maturity, independent input and activity evidence is needed, such as testing capacity, case investigation performance and death-recording processes at zone level, evidence that is distinct from the regularity of the case series itself. We found very little: WHO reports province-level alert-investigation performance, the share of community deaths and 250 unassigned deaths (WHO Regional Office for Africa, 2026), the repository holds a static count of reverse-transcription polymerase chain reaction (RT-PCR) tests by zone, and its zone series for suspected cases and contacts stop at the end of May. This is not evidence of immaturity in any particular zone. It is a lack of evidence that maturity has been reached, and the framework’s evidentiary-discipline principle is to treat that gap as a bound and not to infer maturity from the regularity of the reported counts. Case and death channels need not mature together, and the quantity a_d_/a_c_ in our ascertainment analysis is exactly the difference between them.

For epidemiological maturity, the second requirement is that the parameters of interest be identifiable and stable. The national ratio has plateaued, which is encouraging. The zone-level coefficients are not stable under a moving cut-off (drift ratios above 1), and the reporting-date delay model cannot identify a clinical delay; by the criterion of convergence under rolling-origin evaluation used in the framework, the zone-level parameters have not yet converged. Our conclusion is therefore not that zone ratios are wrong but that they are not yet ready to be read as fatality estimates.

### 5.5. Implications for surveillance design and policy

A readiness test is useful only if it changes what is collected, displayed and decided. Table 5 links each finding to a design action and to an indicator that a ministry or response coordination body could track. The central recommendation is to separate two uses of a zone ratio: as a trigger for investigation, for which it is already adequate, and as a basis for ranking zones, allocating resources or judging care, for which it is not ready.

**Table 5.** From findings to surveillance-design and policy actions.

| Finding | Design or policy action | Indicator to track |
| --- | --- | --- |
| Zone ratios drift by more than their own uncertainty across cut-offs | Publish zone ratios as dated "reported ratios" with case numbers, interval and drift over the last three reports; do | Number of cases; interval width; |
|  | not rank zones on them | drift ratio |
| A 1.25-fold difference in death-to-case ascertainment could create the contrast | Run periodic ascertainment audits by province: retrospective case search in sampled zones and verification of community deaths against confirmed deaths | Estimated $a_c$ and $a_d$ with intervals, by province |
| Deaths not assigned to a zone; most deaths in the community (WHO Regional Office for Africa, 2026) | Assign every death to a zone of residence and report unassigned deaths separately in every SitRep | Share of deaths unassigned; share of community deaths tested |
| Cases and deaths are often reported on the same date | Record dates of onset, confirmation, admission and death, not only report dates | Share of records with each event date; report-to-event lag |
| Static facility maps explain nothing | Collect functional readiness (open, staffed, supplied, used) for treatment centers instead of inventories | Beds occupied; referral time; supply stock-outs |
| No zone-level evidence of testing or investigation performance | Publish zone-level process indicators alongside case counts, with series that continue through the epidemic | Tests per zone; alerts investigated within 24 hours; contacts followed |
| Aggregates cannot identify patient-level fatality | Link patient records (age, sex, viral load, treatment, outcome, last follow-up) under the ethics approval | Share of confirmed cases with a linked outcome |

### 5.6. Limitations

- Surveillance sensitivity and specificity are not measured and cannot be identified from two totals per zone. The ascertainment analysis shows consequences of assumptions, not facts.
- The primary model rests on 41 zones, 14 of them in North Kivu, with leave-one-zone-out estimates ranging from 1.33 to 1.67. Ecological associations at zone level do not describe individuals.
- The estimates are cut-off dependent by construction of the finding, and the epidemic is ongoing. The drift ratio over three cut-offs is a weak signal when calibrated against simulated stable epidemics (Section 4.7); those simulations assume one true proportion per zone, a fixed North Kivu effect and no change in surveillance, and use 20 replicates per scenario, so they give a reference distribution, not a test.
- Epidemic age, recent case share and reporting-date completeness are output-side indicators of the data stream and do not measure maturity or sensitivity.
- Reporting dates are not event dates; no delay from onset or confirmation to death can be estimated.
- National and zone totals differ by 493 deaths; zone-level conclusions assume that unassigned deaths are not concentrated in the zones compared.
- Zone and national figures were transcribed manually from SitRep PDFs into the repository; transcription errors are possible, although no conflicting values remained after name matching. The definition of confirmed deaths is the authors’ interpretation.
- The facility layers are static snapshots (HOT/OpenStreetMap, 5 October 2026; GRID3, 2025). Several candidate predictors were examined (Appendix B) and no multiplicity correction was applied; all but the province contrast are exploratory.

## 6. Conclusions

Reported deaths per confirmed case in the 2026 Bundibugyo epidemic varied widely between health zones, and the size of the differences changed with every cut-off we examined, as it will again. North Kivu zones had higher adjusted odds of a reported death than Ituri zones in the point estimate at every cut-off since 20 July, by a margin that fell from 2.5 on 30 August to 1.5 on 6 October, with the lower credible limit falling from 1.12 on 30 September to 0.99, so that the interval now reached 1. About a 1.25-fold greater completeness of death reporting relative to case reporting would be enough to produce the current contrast, and the data cannot distinguish this from a real difference in outcome. By the readiness test, the zone-level ratio is a signal for investigation, not a measure of fatality or of care. The test needs only published aggregate data and can be repeated at each report; the actions in Table 5, above all ascertainment audits, event-date recording and patient-record linkage, are what would move zone ratios from signal to estimate.

## Declarations

CRediT authorship contribution statement. Johan G. L. Verheyden: Conceptualization, Methodology, Software, Formal analysis, Data curation, Visualization, Writing -original draft. Celestin Nzanzu Mudogo: Validation, Writing -review & editing. Wolfgang Jacquet: Supervision, Writing -review & editing.

## Ethics

This study used only public, aggregate, non-identifiable secondary data, and no ethics approval was required. The authors’ wider research on the 2026 BVD epidemic is covered by approval UNILU/CEM/089/2026 of the Medical Ethics and Research Committee, University of Lubumbashi (5 October 2026).

Declaration of generative AI and AI-assisted technologies in the writing process. The authors used an AI language model (Claude, Anthropic) to assist with code drafting, citation-style conversion, document formatting, and cross-manuscript consistency editing. All data, coding decisions, source excerpts, analytic findings, and interpretive claims originate from the authors’ own research process. All AI-assisted text was reviewed, verified against source material, and approved by the authors before inclusion, and the authors take full responsibility for the content of the publication.

## Data availability

Facility locations are OpenStreetMap data (c) OpenStreetMap contributors, available under the Open Database License, as exported by the Humanitarian OpenStreetMap Team through the Humanitarian Data Exchange (dataset hotosm_cod_health_facilities, snapshot of 5 October 2026). The analysis is based on official DRC INSP situation reports through SitRep 145 (6 October 2026), published under an MIT license at https://github.com/INRB-UMIE/BDBV2026-Data/tree/main/data/insp_sitrep (commit a3250731ee0ed57ebe977c916aec24670082b33a). A source manifest, extraction decisions, analysis-ready derived data, and reproducibility code are deposited on Zenodo (https://doi.org/10.5281/zenodo.23262564), to the extent permitted by source availability. (INRB-UMIE, 2026)

## Acknowledgements

We thank the Institut National de Recherche Biomédicale (INRB), its Unité de Modélisation et Intelligence Épidémiologique (UMIE) and the Institut National de Santé Publique (INSP) for the routine reporting and data curation that made this analysis possible, and the OpenStreetMap contributors and the Humanitarian OpenStreetMap Team for the facility data.

## Abbreviations

BVD: Bundibugyo virus disease
CrI: credible interval
DRC: Democratic Republic of the Congo
INRB: Institut National de Recherche Biomédicale
INSP: Institut National de Santé Publique
OR: odds ratio
RT-PCR: reverse-transcription polymerase chain reaction
SitRep: situation report
UMIE: Unité de Modélisation et Intelligence Épidémiologique
WHO: World Health Organization.

### Appendix A. Reconciliation and zone-level tables

**Table A.1.** Reconciliation of national and zone totals, 6 October 2026.

| Measure | National banner | Sum of zones | Comment |
| --- | --- | --- | --- |
| Confirmed cases | 8,728 | 8,728 | Agree |
| Confirmed deaths | 4,205 | 3,712 | 493 fewer in the zone table |
| Reported ratio | 48.2% | 42.5% | National figure uses the banner |
| Zones with $\geq 10$ / $\geq 20$ cases | - | 41 / 35 | Modeled / descriptive set |

**Table A.2.** Health zones with at least 10 confirmed cases, 6 October 2026 (41 zones).

| Health zone | Province | Confirmed cases | Confirmed deaths | Reported ratio | Days since first case |
| --- | --- | --- | --- | --- | --- |
| Bunia | Ituri | 1790 | 594 | 33% | 142 |
| Rwampara | Ituri | 1118 | 385 | 34% | 145 |
| Nizi | Ituri | 792 | 286 | 36% | 139 |
| Mongbwalu | Ituri | 710 | 337 | 47% | 142 |
| Katwa | North Kivu | 608 | 373 | 61% | 142 |
| Beni | North Kivu | 409 | 268 | 66% | 132 |
| Mangala | Ituri | 366 | 157 | 43% | 132 |
| Nia Nia | Ituri | 314 | 175 | 56% | 116 |
| Lita | Ituri | 295 | 148 | 50% | 132 |
| Butembo | North Kivu | 286 | 180 | 63% | 142 |
| Bambu | Ituri | 210 | 47 | 22% | 140 |
| Mandima | Ituri | 198 | 98 | 49% | 100 |
| Komanda | Ituri | 176 | 107 | 61% | 132 |
| Musienene | North Kivu | 140 | 69 | 49% | 113 |
| Nyankunde | Ituri | 130 | 37 | 28% | 142 |
| Wamba | Haut-Uélé | 119 | 43 | 36% | 89 |
| Isiro | Haut-Uélé | 102 | 44 | 43% | 86 |
| Pawa | Haut-Uélé | 92 | 43 | 47% | 86 |
| Fataki | Ituri | 84 | 31 | 37% | 113 |
| Tchomia | Ituri | 76 | 25 | 33% | 120 |
| Kalunguta | North Kivu | 63 | 22 | 35% | 136 |
| Mabalako | North Kivu | 51 | 20 | 39% | 116 |
| Boma Mangbetu | Haut-Uélé | 43 | 19 | 44% | 86 |
| Mambasa | Ituri | 41 | 17 | 41% | 128 |
| Damas | Ituri | 40 | 9 | 22% | 132 |
| Kilo | Ituri | 38 | 12 | 32% | 139 |
| Kyondo | North Kivu | 37 | 20 | 54% | 135 |
| Vuhovi | North Kivu | 35 | 10 | 29% | 119 |
| Lolwa | Ituri | 35 | 9 | 26% | 99 |
| Oicha | North Kivu | 34 | 16 | 47% | 136 |
| Masereka | North Kivu | 34 | 11 | 32% | 119 |
| Makiso Kisangani | Tshopo | 27 | 11 | 41% | 89 |
| Biena | North Kivu | 23 | 7 | 30% | 42 |
| Lubero | North Kivu | 23 | 11 | 48% | 66 |
| Mutwanga | North Kivu | 20 | 4 | 20% | 46 |
| Drodro | Ituri | 15 | 7 | 47% | 112 |
| Logo | Ituri | 14 | 6 | 43% | 128 |
| Rimba | Ituri | 14 | 5 | 36% | 127 |
| Manguredjipa | North Kivu | 13 | 5 | 38% | 42 |
| Aungba | Ituri | 12 | 5 | 42% | 132 |
| Adja | Ituri | 12 | 1 | 8% | 80 |

**Table A.3.**
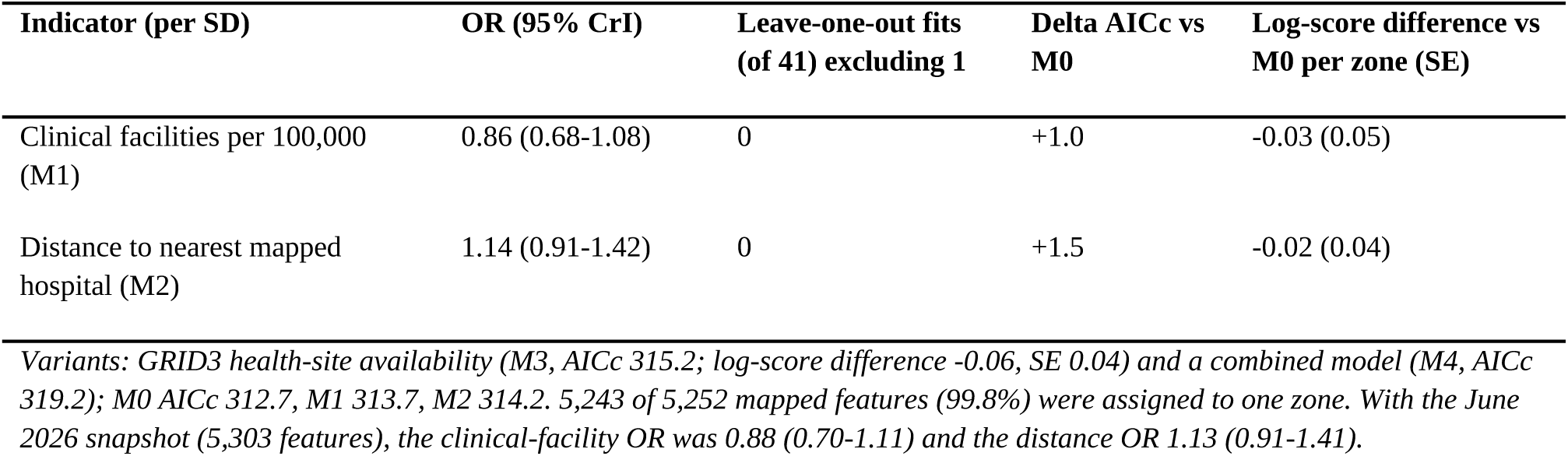
Exploratory facility models (41 zones).

| Indicator (per SD) | OR (95% CrI) | Leave-one-out fits<br>(of 41) excluding 1 | Delta AICc vs<br>M0 | Log-score difference vs<br>M0 per zone (SE) |
| --- | --- | --- | --- | --- |
| Clinical facilities per 100,000<br>(M1) | 0.86 (0.68-1.08) | 0 | +1.0 | -0.03 (0.05) |
| Distance to nearest mapped<br>hospital (M2) | 1.14 (0.91-1.42) | 0 | +1.5 | -0.02 (0.04) |

*Variants: GRID3 health-site availability (M3, AICc 315.2; log-score difference -0.06, SE 0.04) and a combined model (M4, AICc 319.2); M0 AICc 312.7, M1 313.7, M2 314.2. 5,243 of 5,252 mapped features (99.8%) were assigned to one zone. With the June 2026 snapshot (5,303 features), the clinical-facility OR was 0.88 (0.70-1.11) and the distance OR 1.13 (0.91-1.41).*

**Fig. A.1.**
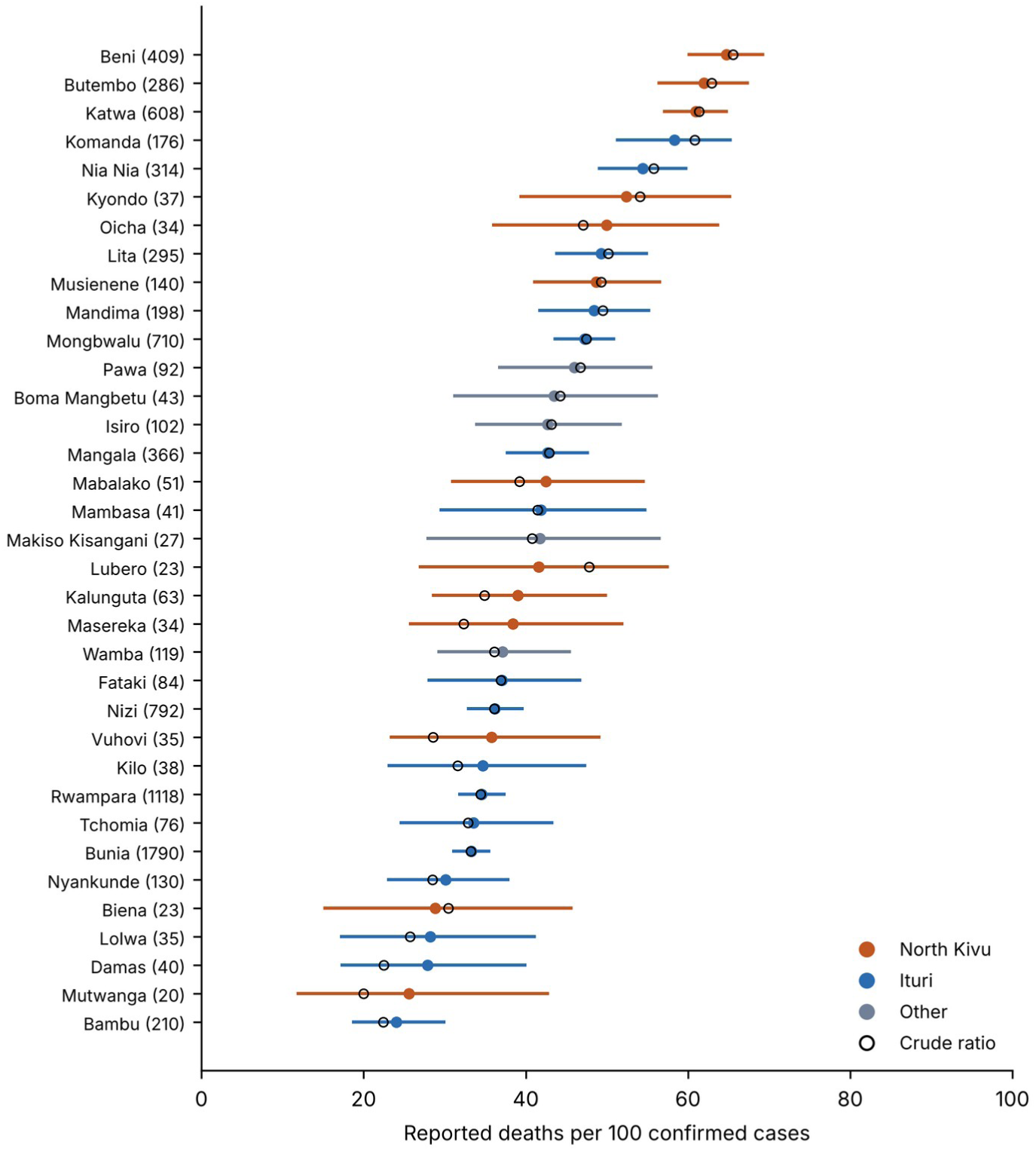
Reported ratio (open circles) and partially pooled estimate with 95% credible interval, by health zone with at least 20 confirmed cases, 6 October 2026. Case counts in parentheses.

**Fig. A.2.**
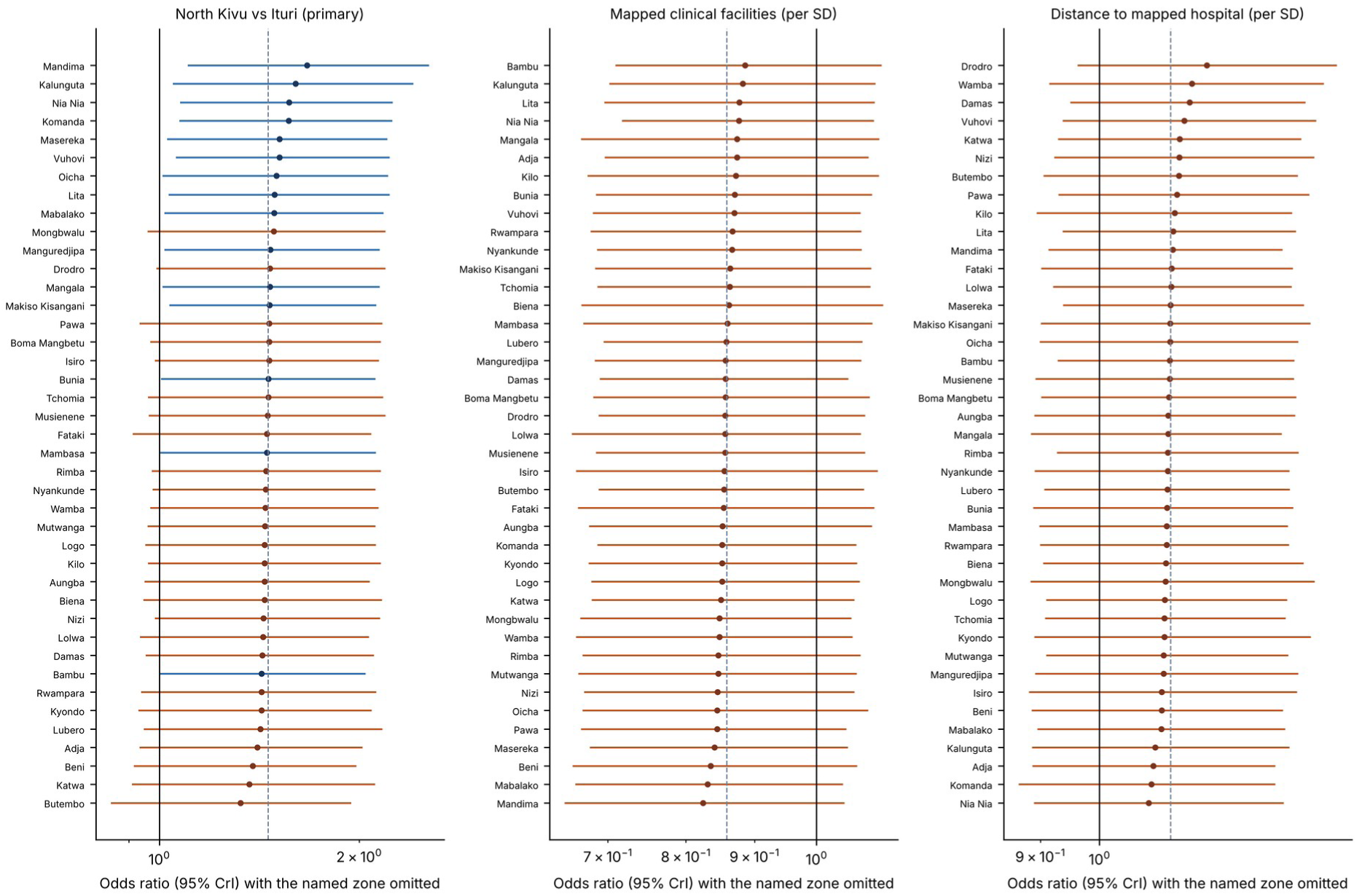
Odds ratio (95% credible interval) when each named zone is omitted in turn: North Kivu versus Ituri (left) and the two facility indicators (center, right). Dashed line: estimate from all zones.

**Fig. A.3.**
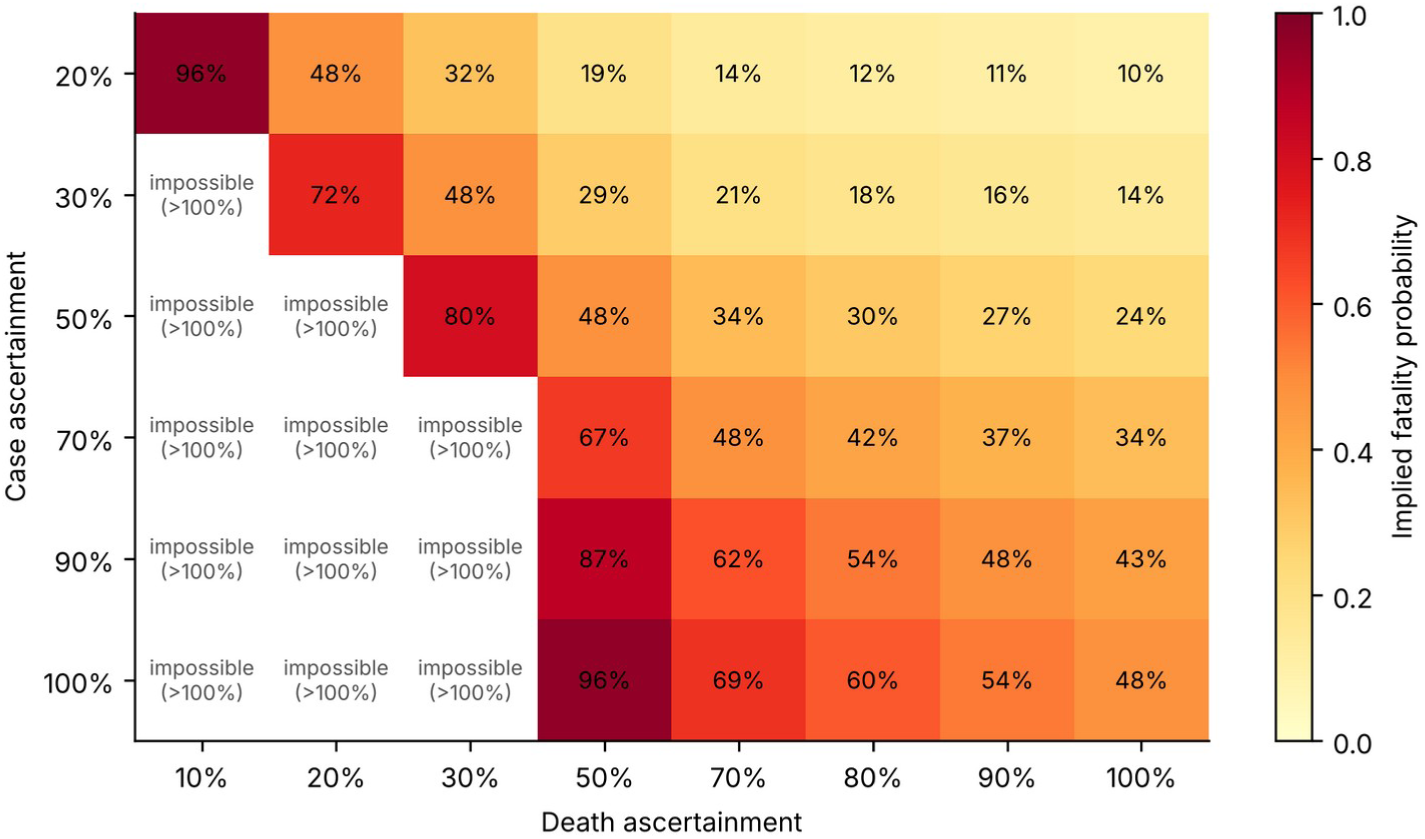
Implied fatality for combinations of case and death ascertainment, given the national reported ratio. Combinations that would imply a fatality above 100% are impossible.

### Appendix B. Candidate predictors and their status relative to the data

No analysis plan was registered or time-stamped before the first analysis, so nothing is described as prespecified. "Fixed" means the model structure was already in the 20 July analysis and was not altered when later data were added.

**Table B.1.**
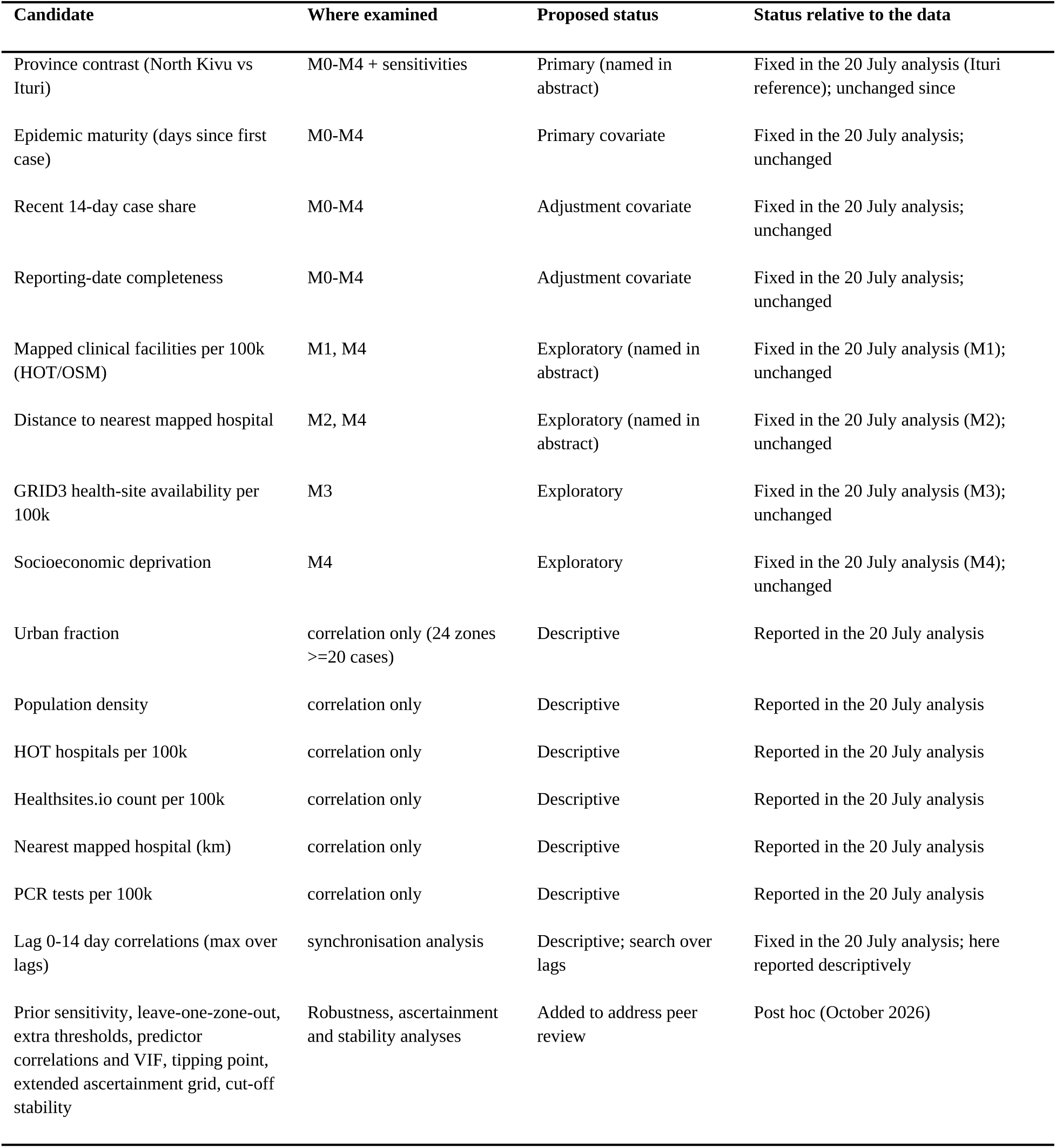
Candidate predictors and analyses.

| Candidate | Where examined | Proposed status | Status relative to the data |
| --- | --- | --- | --- |
| Province contrast (North Kivu vs Ituri) | M0-M4 + sensitivities | Primary (named in abstract) | Fixed in the 20 July analysis (Ituri reference); unchanged since |
| Epidemic maturity (days since first case) | M0-M4 | Primary covariate | Fixed in the 20 July analysis; unchanged |
| Recent 14-day case share | M0-M4 | Adjustment covariate | Fixed in the 20 July analysis; unchanged |
| Reporting-date completeness | M0-M4 | Adjustment covariate | Fixed in the 20 July analysis; unchanged |
| Mapped clinical facilities per 100k (HOT/OSM) | M1, M4 | Exploratory (named in abstract) | Fixed in the 20 July analysis (M1); unchanged |
| Distance to nearest mapped hospital | M2, M4 | Exploratory (named in abstract) | Fixed in the 20 July analysis (M2); unchanged |
| GRID3 health-site availability per 100k | M3 | Exploratory | Fixed in the 20 July analysis (M3); unchanged |
| Socioeconomic deprivation | M4 | Exploratory | Fixed in the 20 July analysis (M4); unchanged |
| Urban fraction | correlation only (24 zones >=20 cases) | Descriptive | Reported in the 20 July analysis |
| Population density | correlation only | Descriptive | Reported in the 20 July analysis |
| HOT hospitals per 100k | correlation only | Descriptive | Reported in the 20 July analysis |
| Healthsites.io count per 100k | correlation only | Descriptive | Reported in the 20 July analysis |
| Nearest mapped hospital (km) | correlation only | Descriptive | Reported in the 20 July analysis |
| PCR tests per 100k | correlation only | Descriptive | Reported in the 20 July analysis |
| Lag 0-14 day correlations (max over lags) | synchronisation analysis | Descriptive; search over lags | Fixed in the 20 July analysis; here reported descriptively |
| Prior sensitivity, leave-one-zone-out, extra thresholds, predictor correlations and VIF, tipping point, extended ascertainment grid, cut-off stability | Robustness, ascertainment and stability analyses | Added to address peer review | Post hoc (October 2026) |

### Appendix C. Reporting-date delay diagnostic

A negative-binomial model of daily national deaths driven by earlier case increments, with a kernel placing mass at lag 0 and a gamma distribution over lags 1 to 21, fitted equally well with and without a same-day reporting-shock term (AICc 990.6 versus 992.2). The free fit gave 63.0% same-day mass and a positive-lag mean of 20.0 days (the upper bound of the search); with the shock term, 57.1% and 20.0 days. The best fixed kernel had same-day mass 0.50 and a lag mean of 14 days. Applying the fitted kernel to zones gave the highest delay-adjusted medians in Beni, Butembo, Katwa, Komanda (74%, 65%, 64%, 63%). These are diagnostics of identifiability, not fatality estimates. Where the delay-weighted exposure (the effective number of cases expected to have resolved) was smaller than the reported deaths, the effective denominator was set equal to the deaths; no modeled zone required this at 30 August, 30 September, 5 October or 6 October.

### Appendix D. Influence of single health zones

**Table D.1.** Leave-one-zone-out summary for key coefficients (41 zones).

| Model | Parameter | Full OR | Min | Max | Fits | Fits excluding 1 | Most influential zone |
| --- | --- | --- | --- | --- | --- | --- | --- |
| M0 | North Kivu | 1.46 | 1.33 | 1.67 | 41 | 15 | Mandima |
| M0 | maturity_z | 1.29 | 1.21 | 1.35 | 41 | 39 | Mutwanga |
| M0 | reporting_z | 0.91 | 0.82 | 0.95 | 41 | 0 | Manguredjipa |
| M1 | North Kivu | 1.78 | 1.63 | 2.21 | 41 | 39 | Mandima |
| M1 | hot_clinical_pc_z | 0.86 | 0.82 | 0.89 | 41 | 0 | Mandima |
| M2 | North Kivu | 1.77 | 1.62 | 1.99 | 41 | 37 | Drodro |
| M2 | nearest_hospital_z | 1.14 | 1.09 | 1.21 | 41 | 0 | Drodro |

### Appendix E. Conflict context and literature check

Conflict appraisal (qualitative judgment; refreshed 8 October 2026). Four explanations of the North Kivu-Ituri difference were weighed qualitatively against source credibility, geographic and temporal fit, mechanism, historical analogy and counter-evidence: contemporaneous violence in North Kivu; direct armed-group administration; chronic structural isolation, disrupted referral and distrust in the Beni-Butembo corridor; and conflict explaining all of the contrast. This appraisal is a qualitative judgment, not a measurement or a statistical analysis: its labels (very low, low, moderate) are not probabilities, and it did not enter any model. No health-zone conflict covariate exists in the repository (its ACLED extract is province-level and ends in May 2026). Attacks on the response have been documented mainly in Ituri (media reports of 12 or more by 18 July, including the Nyakunde hospital and treatment center on 15 July); chronic isolation remained the best-supported conflict-related hypothesis. WHO states that insecurity impedes surveillance, case investigation and contact tracing, and that the North Kivu ratio is under investigation (WHO Regional Office for Africa, 2026).

Literature check (refreshed 8 October 2026). PubMed, publisher sites, WHO Disease Outbreak News and WHO Regional Office for Africa situation reports were searched, first on 24 July 2026 and again on 8 October 2026; this was not a systematic review. Primary sources were read directly; news items and preprints were used for context only. No source was found that estimates the surveillance sensitivity of case or death reporting in the 2026 outbreak by health zone.

### Appendix F. Reproducibility and sampler check

The pipeline comprises a configuration file (pinned repository commit and paths), one script per step (zone table; facility context; models, robustness, ascertainment, AICc and log score; same-day reporting; delay diagnostics; stability across cut-offs; figures; maps), and the model code. Seeds are the cut-off date. Regression tests: at the 30 August 2026 cut-off the zone, facility, same-day and delay steps reproduce the outputs of the earlier analysis exactly, and the model fits reproduce earlier results to within Monte Carlo error. The primary model was refitted with the No-U-Turn sampler (NumPyro) as an independent check, because the earlier package used the same Metropolis class. At 6 October the odds ratios agree with the Metropolis fit to within 0.02, R-hat is 1.00, the effective sample size exceeds 12,000 and no transitions diverged. Two further seeds at 6 October and one run at the 30 September cut-off gave the same agreement (Table F.2).

**Table F.1.**
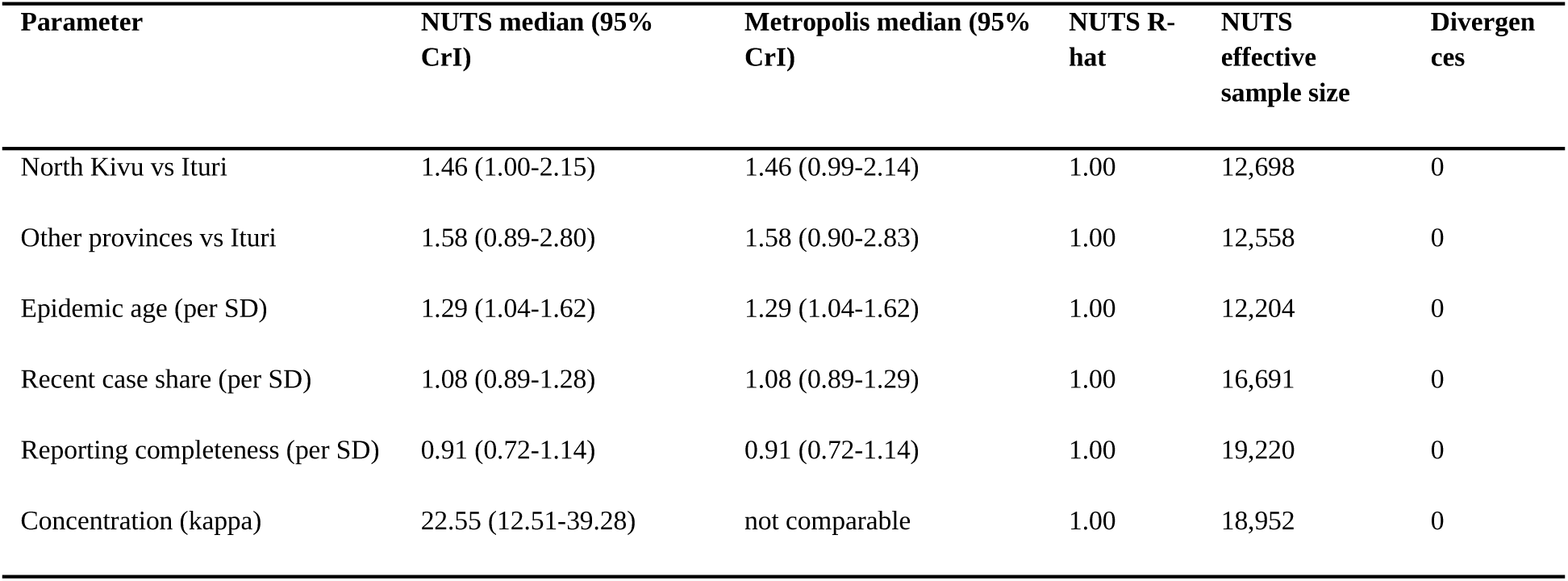
Metropolis versus No-U-Turn (NUTS) fit of the primary model, 6 October 2026 (posterior median and 95% credible interval; odds ratios except kappa).

**Table F.2.**
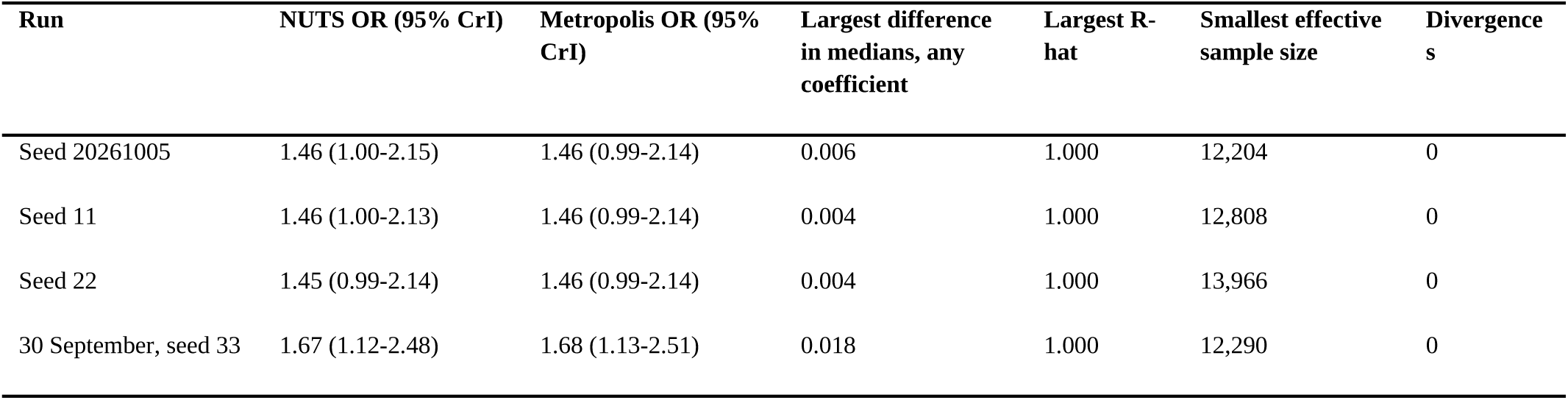
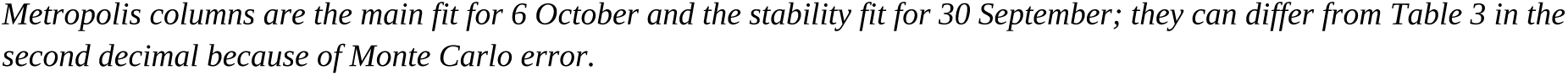
North Kivu odds ratio from repeated No-U-Turn runs (6 October 2026 unless stated).

| Run | NUTS OR (95% CrI) | Metropolis OR (95% CrI) | Largest difference in medians, any coefficient | Largest R-hat | Smallest effective sample size | Divergences |
| --- | --- | --- | --- | --- | --- | --- |
| Seed 20261005 | 1.46 (1.00-2.15) | 1.46 (0.99-2.14) | 0.006 | 1.000 | 12,204 | 0 |
| Seed 11 | 1.46 (1.00-2.13) | 1.46 (0.99-2.14) | 0.004 | 1.000 | 12,808 | 0 |
| Seed 22 | 1.45 (0.99-2.14) | 1.46 (0.99-2.14) | 0.004 | 1.000 | 13,966 | 0 |
| 30 September, seed 33 | 1.67 (1.12-2.48) | 1.68 (1.13-2.51) | 0.018 | 1.000 | 12,290 | 0 |

### Appendix G. Glossary

**Table G.1.** Terms used in the paper.

| Term | Plain meaning |
| --- | --- |
| Reported ratio | Deaths reported divided by confirmed cases reported, in a zone at a given date. Not the chance that a patient dies. |
| Odds / odds ratio (OR) | Odds are deaths per case not (yet) reported dead; an OR compares the odds between two groups. OR 1 = no difference. |
| Credible interval (CrI) | Range of values that, given the model and data, contains the true value with 95% probability. |
| Beta-binomial model | Counts of deaths among cases, allowing true proportions to differ between zones by more than chance alone would produce. |
| Partial pooling / shrinkage | Estimates for small zones are pulled toward the overall pattern, so they are not driven by a handful of cases. |
| Prior | What the model assumes before seeing the data; results are checked under weaker and stronger priors. |
| MCMC, R-hat | Simulation method used to fit the model, and its check that independent runs agree. |
| Leave-one-zone-out | Refit the model omitting each zone in turn to see whether any single zone drives the result. |
| AICc, log score | Measures of how well a model predicts; used to compare models with and without extra predictors. |
| VIF | Variance inflation factor; how much a predictor overlaps with the others. |
| Ascertainment ($a_c$, $a_d$) | The fraction of true cases ($a_c$) or true deaths ($a_d$) that is found and reported. The reported ratio is distorted by $a_d/a_c$. |
| Tipping point | The size of $a_d/a_c$ difference between two provinces that would by itself produce the observed difference in reported ratios. |
| Epidemic age | Days since the zone's first confirmed case. |
| Surveillance maturity / epidemiological maturity | Whether surveillance capacity is adequate to produce a stable record, and whether enough outcomes have accrued for fatality to be estimated (see Introduction). |

### Appendix H. Weekly refits used to calibrate the drift ratio

**Table H.1.** North Kivu odds ratio at weekly cut-offs (phase ending 6 October) and drift ratio over three successive weekly cut-offs.

| Cut-off | Zones | North Kivu OR (95% CrI) | Drift ratio |
| --- | --- | --- | --- |
| 21 July 2026 | 21 | 1.77 (0.91-3.30) | - |
| 28 July 2026 | 24 | 2.83 (1.48-5.40) | - |
| 4 August 2026 | 29 | 2.47 (1.31-4.65) | 1.45 |
| 11 August 2026 | 31 | 2.63 (1.41-4.84) | 0.43 |
| 18 August 2026 | 31 | 2.58 (1.46-4.58) | 0.22 |
| 25 August 2026 | 32 | 2.81 (1.63-4.79) | 0.31 |
| 1 September 2026 | 33 | 2.29 (1.38-3.73) | 0.80 |
| 8 September 2026 | 36 | 1.76 (1.10-2.68) | 2.06 |
| 15 September 2026 | 39 | 2.09 (1.15-3.81) | 0.87 |
| 22 September 2026 | 41 | 2.15 (1.32-3.53) | 0.80 |
| 29 September 2026 | 41 | 1.61 (1.07-2.41) | 1.39 |
| 6 October 2026 | 41 | 1.46 (0.98-2.17) | 1.90 |

*Weekly refits used 15,000 draws per chain, so values for 6 October differ slightly from Tables 2 and 3. Phases ending 3 October and 30 September are in the output file R13\_drift\_real\_weekly.csv of the reproducibility package.*

## References

Akilimali, P., Ebengo, D. M., Scarpino, S. V., Amuri-Aziza, A., Wawina-Bokalanga, T., Matondo-Mbundu, P., Kanku, B., Kinganda-Lusamaki, E., Makangara, J.-C., Mukadi-Bamuleka, D., Rojek, A., Kraemer, M. U. G., Ngandu, C., Mwamba, D., & Mbala-Kingebeni, P. (2026). Clinical characteristics of patients infected with Bundibugyo virus, DRC 2026. The New England Journal of Medicine, 395(6), 610–612. 10.1056/NEJMc2608070

Armed Conflict Location & Event Data. (2026). Africa overview: June 2026. Democratic Republic of Congo: The ADF targets civilians in Ituri amidst the Ebola outbreak. https://acleddata.com/update/africa-overview-june-2026

Bah, E. I., Lamah, M.-C., Fletcher, T., Jacob, S., Brett-Major, D. M., Sall, A. A., Shindo, N., Fischer, W. A., Lamontagne, F., Sow, M. S., Bausch, D. G., Barry, M., Jagatic, T., Sprecher, A., Lawler, J. V., Mayet, T., Jacquerioz, F. A., Méndez Baggi, M. F., Vallenas, C., Clement, C., et al. (2015). Clinical presentation of patients with Ebola virus disease in Conakry, Guinea. The New England Journal of Medicine, 372(1), 40–47. 10.1056/NEJMoa1411249

Bastos, L. S., Economou, T., Gomes, M. F. C., Villela, D. A. M., Coelho, F. C., Cruz, O. G., Stoner, O., Bailey, T., & Codeço, C. T. (2019). A modelling approach for correcting reporting delays in disease surveillance data. Statistics in Medicine, 38(22), 4363–4377. 10.1002/sim.8303

Bhadelia, N. (2026). Bundibugyo virus outbreak: When a concerning pathogen meets a humanitarian emergency. Nature Reviews Microbiology, 24(8), 515–516. 10.1038/s41579-026-01332-9

Doshi, R. H., Garbern, S. C., Kulkarni, S., Perera, S. M., Fleming, M. K., Muhayangabo, R. F., Ombeni, A. B., Tchoualeu, D. D., Kallay, R., Song, E., Powell, J., Gainey, M., Glenn, B., Mutumwa, R. M., Mustafa, S. H. B., Earle-Richardson, G., Gao, H., Abad, N., Soke, G. N., Fitter, D. L., et al. (2023). Ebola vaccine uptake and attitudes among healthcare workers in North Kivu, Democratic Republic of the Congo, 2021. Frontiers in Public Health, 11, Article 1080700. 10.3389/fpubh.2023.1080700

Fitzpatrick, G., Vogt, F., Gbabai, O. B. M., Decroo, T., Keane, M., De Clerck, H., Grolla, A., Brechard, R., Stinson, K., & Van Herp, M. (2015). The contribution of Ebola viral load at admission and other patient characteristics to mortality in a Médecins Sans Frontières Ebola case management centre, Kailahun, Sierra Leone, June– October 2014. The Journal of Infectious Diseases, 212(11), 1752–1758. 10.1093/infdis/jiv304

Forna, A., Dorigatti, I., Nouvellet, P., & Donnelly, C. A. (2020a). Spatiotemporal variability in case fatality ratios for the 2013–2016 Ebola epidemic in West Africa. International Journal of Infectious Diseases, 93, 48–55. 10.1016/j.ijid.2020.01.046

Forna, A., Dorigatti, I., Nouvellet, P., & Donnelly, C. A. (2021). Comparison of machine learning methods for estimating case fatality ratios: An Ebola outbreak simulation study. PLoS ONE, 16(9), Article e0257005. 10.1371/journal.pone.0257005

Forna, A., Nouvellet, P., Dorigatti, I., & Donnelly, C. A. (2020b). Case fatality ratio estimates for the 2013–2016 West African Ebola epidemic: Application of boosted regression trees for imputation. Clinical Infectious Diseases, 70(12), 2476–2483. 10.1093/cid/ciz678

Garske, T., Cori, A., Ariyarajah, A., Blake, I. M., Dorigatti, I., Eckmanns, T., Fraser, C., Hinsley, W., Jombart, T., Mills, H. L., Nedjati-Gilani, G., Newton, E., Nouvellet, P., Perkins, D., Riley, S., Schumacher, D., Shah, A., Van Kerkhove, M. D., Dye, C., & Ferguson, N. M. (2017). Heterogeneities in the case fatality ratio in the West African Ebola outbreak 2013–2016. Philosophical Transactions of the Royal Society B: Biological Sciences, 372, Article 20160308. 10.1098/rstb.2016.0308

Ghani, A. C., Donnelly, C. A., Cox, D. R., Griffin, J. T., Fraser, C., Lam, T. H., Ho, L. M., Chan, W. S., Anderson, R. M., Hedley, A. J., & Leung, G. M. (2005). Methods for estimating the case fatality ratio for a novel, emerging infectious disease. American Journal of Epidemiology, 162(5), 479–486. 10.1093/aje/kwi230

Ilunga Kalenga, O., Moeti, M., Sparrow, A., Nguyen, V.-K., Lucey, D., & Ghebreyesus, T. A. (2019). The ongoing Ebola epidemic in the Democratic Republic of Congo, 2018–2019. The New England Journal of Medicine, 381(4), 373–383. 10.1056/NEJMsr1904253

[dataset] Institut National de Recherche Biomédicale, & Unité de Modélisation et Intelligence Épidémiologique. (2026). BDBV2026-Data: Processed national and health-zone situation-report data (commit a3250731ee0ed57ebe977c916aec24670082b33a) [Data set]. GitHub. https://github.com/INRB-UMIE/BDBV2026-Data

Institut National de Santé Publique. (2026). SitRep MVE No. 014/2026: Situation épidémiologique de la maladie à virus Ebola (data date 28 May 2026; published 29 May 2026). INSP, Democratic Republic of the Congo. https://github.com/INRB-UMIE/BDBV2026-Data/tree/main/data/insp_sitrep/raw

Izudi, J., & Bajunirwe, F. (2024). Case fatality rate for Ebola disease, 1976–2022: A meta-analysis of global data. Journal of Infection and Public Health, 17(1), 25–34. 10.1016/j.jiph.2023.10.020

Kihanduka, E., Tague, C., Nadarajah, S., Rugendabanga, E., Akilimali, A., Muyisa, B., Mugenyi, N., & Byakika-Kibwika, P. (2026). Bundibugyo ebolavirus emergence in eastern Democratic Republic of Congo: The critical role of risk communication and community engagement in strengthening health security in fragile and conflict-affected settings. Journal of Virus Eradication, 12(3), Article 100633. 10.1016/j.jve.2026.100633

Kratz, T., Roddy, P., Tshomba Oloma, A., Jeffs, B., Pou Ciruelo, D., de la Rosa, O., & Borchert, M. (2015). Ebola virus disease outbreak in Isiro, Democratic Republic of the Congo, 2012: Signs and symptoms, management and outcomes. PLoS ONE, 10(6), Article e0129333. 10.1371/journal.pone.0129333

Kyomba, G. K., Law, M. R., Grépin, K. A., Mayaka, S. M., Mambu, T. N. M., Mbunga, B. K., Hategeka, C., Mapatano, M. A., Konde, J. N. N., Ngo-Bebe, D., Babakazo, P. D., Mafuta, E. M., & Kiyombo, G. M. (2024). Barriers and facilitators to healthcare facility utilization by non-Ebola patients during the 2018–2020 Ebola outbreak in the Democratic Republic of Congo. Global Health Research and Policy, 9(1), 47. 10.1186/s41256-024-00387-6

Lamontagne, F., Fowler, R. A., Adhikari, N. K., Murthy, S., Brett-Major, D. M., Jacobs, M., Uyeki, T. M., Vallenas, C., Norris, S. L., Fischer, W. A., Fletcher, T., Levine, A. C., Reed, P., Bausch, D. G., Gove, S., Hall, A., Shepherd, S., Siemieniuk, R. A., Lamah, M. C., Kamara, R., et al. (2018). Evidence-based guidelines for supportive care of patients with Ebola virus disease. The Lancet, 391, 700–708. 10.1016/S0140-6736(17)31795-6

Lupande Mwenebitu, D., Areda, D., Adroba, P., Mwamba, F., Boribo Kikunda, P., Mukuku, O., Bagenda, D., Wayengera, M., Katoto, P. D. M., & Ngaboyeka, G. (2026). Rapid geographic expansion, decelerating growth, and six-month scenario projections of Bundibugyo virus disease in the Democratic Republic of the Congo: Analysis of situation reports 1–61 [Preprint]. Preprints.org, 202608.1151 (v1). Not peer reviewed.

MacNeil, A., Farnon, E. C., Wamala, J., Okware, S., Cannon, D. L., Reed, Z., Towner, J. S., Tappero, J. W., Lutwama, J., Downing, R., Nichol, S. T., Ksiazek, T. G., & Rollin, P. E. (2010). Proportion of deaths and clinical features in Bundibugyo Ebola virus infection, Uganda. Emerging Infectious Diseases, 16(12), 1969– 1972. 10.3201/eid1612.100627

McGough, S. F., Johansson, M. A., Lipsitch, M., & Menzies, N. A. (2020). Nowcasting by Bayesian smoothing: A flexible, generalizable model for real-time epidemic tracking. PLOS Computational Biology, 16(4), Article e1007735. 10.1371/journal.pcbi.1007735

Mulangu, S., Dodd, L. E., Davey, R. T., Jr, Mbaya, O. T., Proschan, M., Mukadi, D., Manzo, M. L., Nzolo, D., Oloma, A. T., Ibanda, A., Ali, R., Coulibaly, S., Levine, A. C., Grais, R., Diaz, J., Lane, H. C., Muyembe-Tamfum, J. J., Sivahera, B., Camara, M., Kojan, R., et al. (2019). A randomized, controlled trial of Ebola virus disease therapeutics. The New England Journal of Medicine, 381(24), 2293–2303. 10.1056/NEJMoa1910993

Médecins Sans Frontières. (2026, July 16). MSF warns against DRC North Kivu’s exclusion from the upcoming Global Fund malaria funding cycle. https://prezly.msf.org.uk/msf-warns-against-drc-north-kivus-exclusion-from-upcoming-global-funds-malaria-funding-cycle

Roddy, P., Howard, N., Van Kerkhove, M. D., Lutwama, J., Wamala, J., Yoti, Z., Colebunders, R., Palma, P. P., Sterk, E., Jeffs, B., Van Herp, M., & Borchert, M. (2012). Clinical manifestations and case management of Ebola haemorrhagic fever caused by a newly identified virus strain, Bundibugyo, Uganda, 2007–2008. PLoS ONE, 7(12), Article e52986. 10.1371/journal.pone.0052986

Rosello, A., Mossoko, M., Flasche, S., Van Hoek, A. J., Mbala, P., Camacho, A., Funk, S., Kucharski, A., Kebela Ilunga, B., Edmunds, W. J., Piot, P., Baguelin, M., & Muyembe Tamfum, J.-J. (2015). Ebola virus disease in the Democratic Republic of the Congo, 1976–2014. eLife, 4, Article e09015. 10.7554/eLife.09015

Skrable, K., Roshania, R., Mallow, M., Wolfman, V., Siakor, M., & Levine, A. C. (2017). The natural history of acute Ebola virus disease among patients managed in five Ebola treatment units in West Africa: A retrospective cohort study. PLOS Neglected Tropical Diseases, 11(7), Article e0005700. 10.1371/journal.pntd.0005700

Sullivan, N. J. (2026). Bundibugyo virus disease in 2026: Clinical and public health responses. The New England Journal of Medicine, 395(3), 278–289. 10.1056/NEJMra2607216

Van Kerkhove, M. D., Bento, A. I., Mills, H. L., Ferguson, N. M., & Donnelly, C. A. (2015). A review of epidemiological parameters from Ebola outbreaks to inform early public health decision-making. Scientific Data, 2(1), Article 150019. 10.1038/sdata.2015.19

Verheyden, J. G. L., & Nzanzu Mudogo, C. (2026). Surveillance maturity and epidemiological maturity: A staged framework for data readiness in real-time outbreak response [Preprint]. SSRN. 10.2139/ssrn.7496541

Wamala, J. F., Lukwago, L., Malimbo, M., Nguku, P., Yoti, Z., Musenero, M., Amone, J., Mbabazi, W., Nanyunja, M., Zaramba, S., Opio, A., Lutwama, J. J., Talisuna, A. O., & Okware, S. I. (2010). Ebola hemorrhagic fever associated with novel virus strain, Uganda, 2007–2008. Emerging Infectious Diseases, 16(7), 1087–1092. 10.3201/eid1607.091525

WHO Ebola Response Team (2014). Ebola virus disease in West Africa — the first 9 months of the epidemic and forward projections. The New England Journal of Medicine, 371, 1481–1495. 10.1056/NEJMoa1411100

World Health Organization Regional Office for Africa. (2026). Bundibugyo virus disease outbreak, Democratic Republic of the Congo: Weekly external situation report 15 (data to 23 August 2026). https://www.afro.who.int/sites/default/files/2026-08/Wk_Ext_SitRep_1523082026_0.pdf

World Health Organization. (2026a, May 29). Ebola disease caused by Bundibugyo virus, Democratic Republic of the Congo and Uganda (Disease Outbreak News).https://www.who.int/emergencies/disease-outbreak-news/item/2026-DON605

World Health Organization. (2026b, June 8). Ebola disease caused by Bundibugyo virus, Democratic Republic of the Congo and Uganda (Disease Outbreak News).https://www.who.int/emergencies/disease-outbreak-news/item/2026-DON606

World Health Organization. (2026c, July 3). Ebola disease caused by Bundibugyo virus, Democratic Republic of the Congo and Uganda (Disease Outbreak News).https://www.who.int/emergencies/disease-outbreak-news/item/2026-DON612

World Health Organization. (2026d, July 17). Ebola disease caused by Bundibugyo virus, Democratic Republic of the Congo and Uganda (Disease Outbreak News).https://www.who.int/emergencies/disease-outbreak-news/item/2026-DON613

World Health Organization. (2026e, September 10). Ebola disease caused by Bundibugyo virus, Democratic Republic of the Congo (Disease Outbreak News).https://www.who.int/emergencies/disease-outbreak-news/item/2026-DON617

World Health Organization. (2026f, May 25). WHO Director-General’s remarks at the virtual ministerial briefing on the Bundibugyo Ebola outbreak [Speech]. https://www.who.int/news-room/speeches/item/who-director-general-s-remarks-at-the-virtual-ministerial-briefing-on-the-bundibugyo-ebola-outbreak-25-may-2026

